# Opening up safely: public health system requirements for ongoing COVID-19 management based on evaluation of Australia’s surveillance system performance

**DOI:** 10.1101/2021.12.06.21266926

**Authors:** Kamalini Lokuge, Katina D’Onise, Emily Banks, Tatum Street, Sydney Jantos, Mohana Baptista, Kathryn Glass

## Abstract

**Background:** Ongoing management of COVID-19 requires an evidence-based understanding of the performance of public health measures to date, and application of this evidence to evolving response objectives. This paper aims to define system requirements for COVID-19 management under future transmission and response scenarios, based on surveillance system performance to date.

**Methods:** From 1^st^ November 2020 to 30^th^ June 2021 community transmission was eliminated in Australia, allowing investigation of system performance in detecting novel outbreaks, including against variants of concern (VoCs). We characterised surveillance systems in place from peer-reviewed and publicly available data, analysed the epidemiological characteristics of novel outbreaks over this period, and assessed surveillance system sensitivity and timeliness in outbreak detection. These findings were integrated with analysis of other critical COVID-19 public health measures to establish requirements for future COVID-19 management.

**Findings:** Australia reported 25 epidemiologically distinct outbreaks and 5 distinct clusters of cases in the study period, all linked through genomic sequencing to breaches in quarantine facilities housing international travellers. Most (21/30, 70%) were detected through testing of those with acute respiratory illness in the community, and 9 through quarantine screening. For the 21 detected in the community, the testing rate (percent of the total State population tested in the week preceding detection) was 2.07% on average, was higher for those detected while prior outbreaks were ongoing. For 17/30 with data, the delay from the primary case to detection of the index case was, on average 4.9 days, with 10 of the 17 outbreaks detected within 5 days and 3 detected after > 7days. One outbreak was preceded by an unexpected positive wastewater detection. Of the 24 outbreaks in 2021, 20 had publicly available sequencing data, all of which were VoCs. Surveillance for future VoCs using a similar strategy to that used for detecting SARS-CoV-2 to date would necessitate a 100-1,000-fold increase in capacity for genomic sequencing.

**Interpretation:** Australia’s surveillance systems performed well in detecting novel introduction of SARS-CoV-2 in a period when community transmission was eliminated, introductions were infrequent and case numbers were low. Detection relied on community surveillance in symptomatic members of the general population and quarantine screening, supported by comprehensive genomic sequencing. Once vaccine coverage is maximised, the priority for future COVID-19 control will shift to detection of SARS-CoV-2 Vos associated with increased severity of disease in the vaccinated and vaccine ineligible. This will require ongoing investment in maintaining surveillance systems and testing of all international arrivals, alongside greatly increased genomic sequencing capacity. Other essential requirements for managing voices are maintaining outbreak response capacity and developing capacity to rapidly engineer, manufacture, and distribute variant vaccines at scale. The most important factor in management of COVID-19 now and into the future will continue to be how effectively governments support all sectors of the community to engage in control measures.

## Background

Countries recognised as having the best initial responses to the COVID-19 pandemic include Singapore, Taiwan, South Korea, Australia, New Zealand, Canada, Iceland, UAE, Germany, and Greece. These countries were largely successful in minimising morbidity and mortality prior to the availability of vaccines.^1, 2^ Key to this success was early identification of community transmission through surveillance, control of outbreaks through non-pharmaceutical public health measures restricting mobility and social interaction, and effective management of international borders which reduced the risk of subsequent re-introductions.^3, 4^ As well as reducing health impacts, these measures have limited the number and duration of restrictions on community mobility and interaction. Consequently the social and economic burdens of the pandemic were lower in these settings than in many other countries.^5^

In the initial phases of the global pandemic, reliance on nationwide lockdowns in Australia was necessitated by delayed closure of international borders,^6^ which allowed community transmission to establish itself throughout many urban settings, and by limited surveillance, testing, and public health contact tracing capacity for detection and control of such transmission.^7^ From November 2020 to June 2021, with improved capacity and capability of public health systems, and a reduction in the number of introductions from international arrivals, Australia was able to effectively identify and control outbreaks with a combination of measures, which sometimes included short lockdowns.

However, since late 2020, the epidemiology and thus control of COVID-19 has changed due to two critical and opposing developments. The first is the emergence of novel variants of SARS-COV-2 with higher transmissibility.^8^ The second is the availability of several vaccines against SARS-CoV-2,^9^ many of which have been in use since early 2021, one year after the disease began circulating globally.

Current COVID-19 vaccines are effective at limiting symptomatic disease, hospitalisation, and mortality from the virus,^10^ and reduce disease transmission for most variants seen to date.^11^ It is likely that the initial round of vaccine programs in most high-income countries will be completed by 2022.^12^ However, it will take much longer for low-and lower-middle income countries to achieve adequate coverage due to supply constrictions and inequitable global distribution.^13-15^

Broadly, Australia’s vaccination program initially prioritised distribution to those at higher risk of exposure to the virus and those most at risk of severe illness if infected. As vaccine supply increased, the entire population >12 years of age and older became eligible for vaccination. However, the potential for widespread community transmission of SARS-CoV-2 in Australia may remain following completion of the planned vaccination program if herd immunity is not achieved. Opening of international borders in such a context will mean continual introductions of disease.

Furthermore, until transmission is controlled globally, we are likely to continue to see the emergence of new variants, which may have higher transmission potential and increased severity of disease, including in the vaccinated and in vaccine-ineligible. There is therefore a strong imperative to understand the continuing public health system requirements for detecting and controlling community transmission, including of VOCs, once planned vaccine programs are completed and other public health control measures are lifted.

Community transmission of SARS-CoV-2 in Australia was eliminated in the period from 1^st^ November 2020 to 30^th^ June 2021, and all recorded outbreaks in the same period have been linked through epidemiological investigation and genomic sequencing to specific breaches in quarantine facilities that housed returned international travellers. Several breaches resulted in local transmission in the community, but all were subsequently contained, aside from outbreaks in Victoria and NSW that commenced late in the study period and subsequently led to ongoing community transmission from July 2021 onwards. The study period (1^st^ November 2020 to 30^th^ June 2021) provides an ideal context in which to evaluate surveillance performance as community transmission was controlled and case detection was close to complete.^16^

The aim of this paper is therefore to define critical public health requirements for the ongoing management of COVID-19, including surveillance system requirements to support detection and control. This information is critical, including in future contexts of widespread vaccination, reopening of international borders, and the emergence of variants of concern (VoCs). To address this aim, we:

1. Characterised and evaluated the performance of COVID-19 public health surveillance systems in detecting novel outbreaks of COVID-19 community transmission in the period from November 2020 to June 2021 in Australia;
2. Integrated the results of the above surveillance system evaluation with data on other critical public health measures for COVID-19 management, including outbreak response and vaccination; and
3. Used this evidence to define public health system requirements for current and future COVID-19 prevention, detection, and response.

## Methods

### Definitions - see Supplementary Materials (1)

#### Testing rates

were defined as the proportion of the total population in that State tested for Sars-CoV-2. These were calculated for the week preceding the detection of the outbreak, for the State in which the outbreak occurred.

#### Outbreak definition

Outbreaks of COVID-19 included in this analysis included any in which one or more cases were due to community (locally acquired) transmission within the specified time from 1^st^ November 2020 to 30^th^ June 2021. Community transmission included any local transmission in the community, as well as any cases in quarantine or health workers in direct contact with international travellers. It also included international travellers who were infected while in quarantine in Australia, but excluded those international travellers who were infected prior to entering quarantine or who were infected by household/family contacts with whom they were residing while in quarantine. Each individual outbreak was defined as all cases epidemiologically linked to each other. If epidemiological links were not identified between distinct clusters of cases, these clusters were still considered different outbreaks even if a common source in a returned overseas traveller was identified through genomic sequencing.

#### Cluster definition

A cluster was defined as a distinct group of cases initially identified as an outbreak but which was eventually linked to another outbreak via epidemiological evidence and therefore did not meet the above outbreak definition. However, as these clusters were included as they provide useful information on surveillance system performance in detecting transmission.

#### Index case

The index case was defined as the first reported case identified in that outbreak.

#### Source case

The source case was defined as the case with the earliest date of symptom onset and/or infectivity that could be linked to that outbreak through genomic evidence.

#### Primary case

The primary case was defined as the case with the earliest date of symptom onset and/or infectivity that could be epidemiologically linked to that outbreak.

(Note: in general, the epidemiological terms ‘primary’ and ‘source’ case are used interchangeably as they are the same individual. However, in this paper we have differentiated these 2 terms and adopted slightly different definitions, as described above, to account for the fact that the true primary /source case, (which was always an international arrival) was often not linked epidemiologically, despite being linked through genomics. By differentiating, we were able to better describe the characteristics of the outbreaks and their detection).

### Search strategy

We searched peer-reviewed literature and publicly available data to:

a. characterise the surveillance systems in place at the time of novel outbreaks (system characteristics, per capita testing rates, wastewater surveillance sites in operation);
b. quantify and analyse the epidemiological characteristics of outbreaks occurring during this period (index cases, first diagnosed cases, total cases at time of first case being identified, links or transmission routes between cases, time from identification of the initial case to onset of infectivity in infected contacts); and
c. based on these results, conduct a public health analysis to establish requirements for COVID-19 detection and control under future transmission and response scenarios.

### Data sources

The epidemiological data on outbreaks in Australia from 1^st^ November 2020 to 30^th^ June 2021 were collected from a range of government and media sources depending on the location of the outbreak, including State and Federal government websites. When information was not available using government health department resources, online platforms of national public and commercial media outlets were used, supplemented by media sources local to the area of the outbreak.

The data source for state-level COVID-19 testing numbers was www.covid19data.com.au, a volunteer-run not-for-profit online platform. The website verifies data with Federal, State and Territory health departments before publishing. Population data used to calculate state-level COVID-19 testing rates were sourced from publicly available, government-managed data including the Australian Bureau of Statistics (ABS).

#### Surveillance system performance

The key measure of surveillance system performance used were sensitivity and timeliness in detecting introductions of COVID-19 into the community. For the whole surveillance system and for each component of the system, we assessed these outcomes by comparing when and how an outbreak was first detected through surveillance, using outbreak source and seeding information identified through subsequent epidemiological investigation, genomic testing and linkages.

##### Sensitivity

number of new outbreaks in study period first detected through surveillance / total number of new outbreaks in study period.

##### Timeliness

delay between the seeding event linked to the primary (source) case and the date the index case was reported.

#### Public health analysis

Public health analysis was undertaken by defining potential future SARS-CoV-2 transmission and response scenarios based on the pandemic to date. These future scenarios were developed based on the evolution of SARS-CoV-2 transmission and pathogenicity, alongside the development of public health control measures. The implications of these scenarios for surveillance system requirements were then assessed given the results of the surveillance system performance evaluation. Finally, the requirements for other key, non-surveillance-related public health control measures were considered using a similar approach.

## Results

### Components of Australia’s COVID-19 public health surveillance system

The different components of COVID-19 surveillance and screening used for detection of new outbreaks of local transmission are described in the following sections.

#### Community-based surveillance for disease in symptomatic individuals

A key component of Australia’s response has been surveillance for community transmission based on testing those with acute respiratory symptoms. The World Health Organization (WHO) recommended a syndromic screening definition for testing of “fever and cough” early in the outbreak.^17^ Although often reported in patients with COVID-19, a substantial proportion of confirmed COVID-19 cases do not report these symptoms.^18^ Australia has utilised a broader syndromic screening definition to target testing in the community since April 2020, which included any acute respiratory symptoms such as cough, sore throat, runny nose, cold symptoms, or flu-like symptoms.^19^

Currently, collection of most samples for community-based surveillance and testing in Australia is carried out in government-run testing centres and General Practice Respiratory Clinics. Between 30%^20^ and 50% of those in the community with acute respiratory symptoms that meet the screening definition are being tested for COVID-19. Testing rates at an overall population level vary over time and by State, with marked increases in areas with confirmed outbreaks.

#### Genomic surveillance

During the study period, Australia aimed to fully sequence all cases of SARS-CoV-2 infection identified in the country. This includes all cases in returned overseas travellers. AusTrakka, a platform for sharing of sequencing data, supports the Communicable Disease Genomic Network (CDGN) which includes all laboratories conducting SARS-CoV-2 genomics/diagnostics, to share sequencing data to support epidemiological outbreak investigation across the country. As of 7 May 2021, it is estimated that 58% of COVID-19 samples have been sequenced.^21^

#### Screening of residents and staff linked to overseas traveller quarantine

Almost all international arrivals to Australia during the study period were required to undertake two weeks of mandatory supervised quarantine.^22^ During this period, current policy required testing in the first 48 hours and then again between days 10 to 12 of quarantine for SARS-CoV-2 infection,^22^ although exact protocols varied between States. Following incidents of transmission between travellers in quarantine facilities, national policy was changed in June 2021 to require returnees to undergo testing on day 16 (i.e. two days after release from quarantine),^23^ although this was already policy in several States prior to this. Routine, intensive testing of staff working in quarantine facilities commenced in January 2021; prior to this, testing was occurring, but less frequently.

#### Wastewater surveillance

Several states in Australia have ongoing wastewater testing programs for COVID-19 which screen for viral fragments. Positive samples are reported publicly by health departments to encourage those in the catchment areas to present for testing if they have symptoms. Positive wastewater detections are broadly classified as ‘expected’ (known positive or recovered cases in the catchment) or ‘unexpected’.

### Epidemiology of outbreaks, November 2020-June 2021 in Australia

Outbreaks and clusters initiated between 1^st^ November 2020 to 30 June 2021 in Australia are described in *Supplementary Materials (2): Characteristics of community SARS-CoV-2 outbreaks, Australia, November 2020-June 2021*. A total of 30 outbreaks and clusters were reported in the study period. Amongst these, 25 epidemiologically distinct outbreaks (i.e., no epidemiological link identified between that outbreak and others) were identified. All outbreaks were initially detected in major metropolitan areas, aside from one in the Northern Territory detected in a remote mine. Six outbreaks and one cluster occurred in 2020, and the remaining 24 in 2021.

In addition to the above outbreaks, five distinct clusters were identified: the NSW Berala cluster (linked back to a December 2021 case in a quarantine worker): the VIC Coburg cluster (linked to a February 2020 quarantine hotel outbreak); a health worker-related cluster in a QLD hospital in March 2020 (linked to a quarantine-related health care worker infection in the same hospital in the same month); a VIC Port Melbourne cluster in May 2021 (linked through a workplace to another Melbourne outbreak); and a QLD cluster linked to an NT mine outbreak in June 2021.

#### Index case

Of the 25 outbreaks, the index case was detected through community surveillance in 18 (72%) outbreaks, and through quarantine–related screening of staff or residents in 7 (28%). Of those detected through community surveillance, five were detected in individuals who had visited multiple states and/or territories while infectious. One was detected in a family member of a quarantine worker. Of those detected in quarantine residents or workers linked to quarantine, three were detected in residents after they left quarantine, and one was identified through testing of a symptomatic hospital healthcare worker who had treated patients referred from quarantine facilities.

Of the five clusters linked to outbreaks but initially considered distinct, the index cases in these clusters were all detected through community-based surveillance (including one in a healthcare worker), and only following detection of the index case were they then linked back to other outbreaks through previously undetected community transmission.

Of the 24 outbreaks or clusters that occurred in 2021, the index cases in 18 (75%), were detected through community surveillance (including two in healthcare workers), and in 6 (25%) through quarantine screening.

#### Source case

All outbreaks (and related clusters) aside from one were linked through genomic sequencing to breaches in quarantine facilities housing international travellers. The one exception was an outbreak in which the index (also primary) case was an international air crew limousine driver, and health authorities have suggested the most likely source is therefore also international. Sequencing also showed that a single overseas traveller was the common source for NSW-Croydon, VIC-Blackrock, VIC-Vermont, all of which were seeded by the outbreak that commenced in NSW in December 2020 (Northern Beaches).

#### Primary case and early transmission links

Of the 30 outbreaks and clusters, an epidemiological link between the source case and the primary case (including those where both were the same) was identified in 17 (57%). The 13 outbreaks or clusters where no direct link was found included one in South Australia in November 2020; the NSW Northern Beaches outbreak and three related outbreaks or clusters with the same genomic sequence, one in NSW and two in Victoria; an outbreak in NSW in May 2021; and the outbreak suspected to be linked to air crew infecting a driver in NSW; and six outbreaks in Victoria in May and June 2021 all with the same genomic sequence as a returned overseas traveller from South Australian quarantine.

Of the 30 outbreaks and clusters, all except one included transmission in the community outside a quarantine setting. The one exception was an outbreak in family of returned travellers infected while in quarantine. This family were detected as positive while in quarantine but subsequently found to have the same genomic sequence as the source case who was residing in the opposite room during their quarantine period.

#### Community mobility and interaction at time of disease incursion

19 of the 25 outbreaks were seeded (i.e., the transmission event from the primary case) during periods of close to normal mobility (i.e., no masks in use, mass gatherings allowed, and no restrictions on the number of visitors in the home). Of the remaining outbreaks, four of these – although detected during periods in which restrictions were in place – are likely to have been seeded prior to implementation of those restrictions. This included the Croydon and Berala outbreaks detected in late December 2020 in NSW, which were identified during restrictions imposed on the 20^th^ of December following the detection of the Northern Beaches outbreak; and VIC outbreaks in May and June (Arcare and West Melbourne), which were detected during restrictions imposed following the initial clusters detected in Melbourne in late May.

The outbreak in April in WA commenced while mask wearing in public was in place following a previous outbreak, however general community mobility and social interaction were not restricted.

All five clusters occurred during periods when there were other ongoing outbreaks and some level of restrictions.

### Surveillance system performance in detecting outbreaks

#### Community-based disease surveillance and outbreak detection

Initial identification of 21 (70%) of the 30 outbreaks/clusters was through testing of symptomatic community members, including 2 healthcare workers. For these 21 outbreaks/clusters, at the time of detection of the index case, the testing rate (percent of the total State population tested in the week preceding detection) was 2.07% on average. The average testing rate varied over time (1.68% in 2020 and 2.23% in 2021), and by the State in which the outbreak was detected.

In 2020, the testing rates in the week prior to detection of the outbreak in the 5 outbreak/clusters detected through community surveillance ranged from 0.89% to 3.35% of the State population. The three outbreaks/clusters with rates at around 0.9%, in NSW, VIC and SA, were detected in periods without any other ongoing outbreaks; one outbreak and one cluster in NSW both had testing rates of over 3%, both were detected in periods when preceding outbreaks were still active.

In 2021, the testing rates in the week prior to detection of the outbreak in the 20 outbreaks and four clusters ranged from 0.83% to over 4%. There were six outbreaks with testing rates over 3%, including five outbreaks in Victoria from May-June, and the outbreak in NT. The outbreak/clusters in Victoria in 2021 with testing rates of 1-2% were the Port Melbourne cluster and the Whittlesea cluster. These were both the initial outbreak/clusters in the May-June period, with the other outbreaks in this period occurring while these outbreaks were still active. There were two outbreaks/clusters with rates of less than 1%, these were the outbreaks in in Brisbane in March, and the Eastern suburbs outbreak in NSW in June.

#### Screening of quarantine residents and staff

Of the 24 outbreak/clusters that occurred from January 2021 onwards, when intensive regular testing of quarantine staff commenced nationally, seven were identified through screening of quarantine residents or staff, including one case in a resident who had left hotel quarantine and was tested after becoming symptomatic.

#### Genomic surveillance

All outbreaks/clusters were due to viruses with a novel genome sequence and not identified as circulating in the community prior to that outbreak/s, as described previously. Of the 24 outbreak/clusters in 2021, information on the strain of SAR-CoV-2 was available for 20. All were variants of concern. Of these, seven were due to the Delta variant, five to the Kappa variant, seven to the Alpha variant and one was reported as a VoC without further specification.

#### Timeliness of detection

##### Time from primary case to detection of the outbreak

Of the 30 outbreak/clusters, data was available on 17 to allow calculation of this outcome. On average, the delay was 4.9 days (4.7 for outbreaks and 5.7 for clusters), with 10 of the 17 outbreak/clusters detected within five days and three detected after more than a week. These three included the NSW Berala cluster in January 2021 (10 days delay), the West Melbourne outbreak in June 2021 (detected after 9 days) and the Queensland hospital outbreak (10 days delay).

Of the 17 outbreak/clusters with data, 11 were detected through community surveillance, four through quarantine and two in health care workers who became symptomatic. The average delay for these groups was 5.7 days for community detection, 4 days for quarantine detection and 3 days for healthcare worker detections.

#### Wastewater surveillance

All States/catchments reporting outbreaks/clusters in the period from 1^st^ November 2020 to 17 June 2021 except for the NT had wastewater surveillance programs during this period. Others that did not report outbreaks also have surveillance programs (ACT). All these locations reported positive wastewater samples within the study period (data not presented). Of those positive samples that were unexpected (i.e., no known positive or recovered cases in the catchment at the time of detection), one may have preceded the seeding event and therefore helped identify the outbreak through raising awareness and testing levels in the community (Coburg, VIC). The remaining unexpected detections were not linked subsequently to reported outbreaks.

### Surveillance system requirements for current and future control

The following section presents public health analysis undertaken by defining potential future SARS-CoV-2 transmission and response scenarios and analysing their implications for surveillance system requirements. These future scenarios have been developed based on the evolution of SARS-CoV-2 transmission characteristics over the course of the pandemic thus far, alongside the evolution of public health control measures, in particular vaccination.

#### Relationship between vaccination and other control measures

Ignoring differences by age and different vaccine characteristics, we achieve herd immunity if V*VE > 1 - 1/R, where V = proportion vaccinated; VE = vaccine efficacy; R = reproductive number. Even if herd immunity is not achieved, control of transmission can still be maintained if any increase in transmission potential, for example through less stringent restrictions, is offset by gains in control made by increased vaccine coverage (i.e., such that the overall reproductive number remains below 1). Once the vaccination program has achieved its targets, public health responses will be based on the post-vaccination effective reproduction number in the community, with the relationship between different measures continuing as defined above. This may also include booster vaccines to address waning immunity against existing strains.^24^ However, irrespective of whether herd immunity is achieved or not, (and if not achieved, whether control of current SARS-CoV-2 strains is aimed for or not), the capacity to prevent, detect and control the spread of novel VoCs will need to be ongoing while transmission is widespread globally.^25^

The most important factor to consider when assessing novel variants will be indications of increased severity, including in those who are vaccinated, and in those ineligible for vaccination. The majority of transmission of SARS-CoV-2 occurs early in the course of the disease, prior to severe disease, and in those not at risk of severe disease.^26^ These characteristics select for strains with increased transmissibility but may not provide selective pressure against strains with higher mortality. The virus has already demonstrated the capacity to evade vaccine or disease-related immunity and some evidence to date suggests that novel strains may also affect younger age groups, and have increased mortality.^8, 16, 27^ This is particularly important for those groups not eligible for vaccination, of which children are the biggest group. There are reports of increasing mortality in children, including <5yo, from those countries experiencing widespread transmission due to the Delta variant.^28^ However, availability of treatments such as monoclonal antibodies and antivirals may moderate the risk of variants being categorised as ‘of concern’,^29^ if these treatments are effective and available to the whole of the population.

#### Community-based surveillance

Table 1 presents a public health analysis of the implications for community-based surveillance system requirements based on future scenarios. Irrespective of whether the decision is made to allow local endemic transmission or not, the surveillance system will need to maintain the capacity to detect variants with increased severity of disease. The key implication is that future surveillance systems will require significant scale up of genomic sequencing capacity to detect VoCs.

**Table 1:** Community-based surveillance system requirements based on scenarios for COVID-19 epidemiology.

| Scenario | Key epidemiological characteristics/ assumptions | Surveillance system capacity needed to meet requirements | Changes needed to surveillance system |
| --- | --- | --- | --- |
| **A. While COVID-19 vaccination program being rolled out** | Risk of substantial morbidity and mortality requires maintaining current community-based surveillance for disease and screening protocols for returned overseas travellers and staff in quarantine facilities, supported by genomic sequencing. | As per current requirements. i.e., As current surveillance system requirements are influenced by community acute respiratory illness level rather than SARS-CoV-2 rates,<sup>1</sup> capacity requirements will be dictated by levels of ARI in the community. | Nil to maintain performance. However, if performance was improved, this is likely to make control of outbreaks, in particular due to Delta variants, easier to control.<br>Performance could be further improved through improving testing rates (currently <50%) and timeliness in the symptomatic, particularly in communities at risk of low engagement in health interventions, and in high-risk occupations (e.g., health care workers). |
| **B (1) current vaccination program completed; herd immunity achieved; current variants circulating,** | Even if importations increase, transmission will be self-limiting in the general population as the reproductive number will remain below 1 overall. | If herd immunity, or even close to herd immunity is achieved through vaccination, SARS-CoV-2 levels will remain low, and community surveillance capacity will continue to be related to non SARS-CoV-2 ARI rates. | Surveillance capacity will continue depend on background ARI rates. (e.g., 4-6 % of the population per week during the study period<sup>2</sup>, likely to increase once social and mobility restrictions are fully lifted).<br>Focus will need to include communities at risk of low uptake of health interventions, which may experience increased disease circulation and morbidity and mortality of both vaccine coverage and testing uptake is low. |
| **B (2) current vaccination program is** | If the decision is made not to aim for complete control of transmission through public | It is likely that SARS-CoV-2 levels will eventually reach levels similar to other acute respiratory illnesses. If SARS-CoV-2 displaces other | If surveillance capacity needs to increase substantially, alternatives to PCR will be needed. Such alternatives are already in widespread use in developed high-burden settings |

<sup>1</sup> Lokuge K, Banks E, Davis S, et al. Exit strategies: optimising feasible surveillance for detection, elimination, and ongoing prevention of COVID-19 community transmission. *BMC Medicine* 2021; **19**(1): 50.
<sup>2</sup> Grundy E, Slattery P, Buckland G, et al. Scrub Project Wave 4: Australians' Views on Private Gatherings, Remote Working and Getting Tested. 24 June 2020. https://www.behaviourworksaustralia.org/scrub-project-wave-4-australians-views-on-private-gatherings-remote-working-and-getting-tested/

|  |  |  |  |
| --- | --- | --- | --- |
| **completed; herd immunity not achieved, current variants of concern circulating** | health measures, not reaching herd immunity will mean endemic circulation of SARS-CoV-2, with the level of circulation dependant on the impact of the public health measures left in place. | respiratory viruses<sup>3</sup> overall ARI levels will remain constant, and therefore also surveillance requirements. If COVID-19 infections are additive to existing ARI burden, an increase in community surveillance capacity will be required. | such as the UK and USA.<sup>4</sup> Although of lower sensitivity,<sup>5</sup> overall performance will be similar or better than PCR if they facilitate more widespread and frequent screening. Prior analysis has shown that at a population prevalence of 3%, a test with 80% sensitivity that can test all the population exhaustively will miss 24% of cases, whereas test with 99.9% sensitivity, but which can only test a third of the population under surveillance, will miss 66% of cases in the population overall.<sup>6</sup> |
| **C. Emergence of new variants of concern** | Given the characteristics of the virus to date, there is potential for new variants of concern to emerge that result in increased mortality in the vaccinated, and in groups ineligible for vaccination (e.g., children) while transmission is widespread globally.<sup>7</sup> | The first level of surveillance for variants of concern relies on detecting SARS-CoV-2, and therefore the requirements outlined in the sections above apply. In addition, testing would need to include a greatly increased number of international arrivals.<br>The second level of surveillance for early and effective detection of variants will rely on genomic sequencing of either all or an adequate proportion of all detected SARS-CoV-2 in the community and in international arrivals. | To achieve similar performance in detecting variants of concern as current capacity to detect SARS-CoV-2, sequencing capacity will need to be similar to current PCR-testing capacity. i.e., 1-2% of the population<br>This equates to 200,000-400,000 samples per week nationally, given Australia's population of ~21million. This is a >1000-fold increase from current sequencing levels., |

ARI: Acute respiratory illness
<sup>3</sup> MacIntyre R. Vaccination for COVID-19 control and considerations for Australia. *Microbiology Australia* 2021; **42**(1): 30-4.
<sup>4</sup> Crozier A, Rajan S, Buchan I, McKee M. Put to the test: use of rapid testing technologies for covid-19. *BMJ* 2021; **372**: n208.
<sup>5</sup> Crozier A, Rajan S, Buchan I, McKee M. Put to the test: use of rapid testing technologies for covid-19. *BMJ* 2021; **372**: n208.
<sup>6</sup> Lokuge K, Banks E, Davis S, et al. Exit strategies: optimising feasible surveillance for detection, elimination, and ongoing prevention of COVID-19 community transmission. *BMC Medicine* 2021; **19**(1): 50.
<sup>7</sup> World Health Organization. COVID-19 Weekly Epidemiological Update, 2021. https://www.who.int/docs/default-source/coronaviruse/situation-reports/20210622-weekly-epi-update_45.pdf?sfvrsn=f7043e1f_10&download=true

#### Genomic sequencing

A population SARS-CoV-2 testing rate of 1-2% per week has been able to detect introductions of the virus early in transmission. To achieve similar performance in detecting variants of concern, capacity will be needed to sequence all SARS-CoV-2 positive samples, until or unless the number of positive samples reaches 1-2% of the population per week. At which point, sequencing would shift to a randomly selected proportion of positive samples to maintain a rate of 1-2% population sequenced per week. This would require the capacity to sequence around 200,000-400,000 samples per week nationally, given Australia’s population of ∼25 million. This is a 1000-fold increase from current levels, and it is unlikely that full genome sequencing can be scaled up to meet such requirements. Development of technologies that can screen for, rapidly and at scale, specific mutations of particular concern will therefore be of great value (e.g., similar to GenXpert tools in screening for anti-tuberculosis drug resistance mutations).

It is difficult to predict if Australia would reach such high infection rates that 1-2% of the population would test positive each week. Data from the US suggests that ∼30% of the population experienced symptomatic infection from February 2020-May 2021,^30^ approximately one third to one half of a 1-2% weekly positivity rate. This US data is prior to circulation of the Delta variant and included periods of restricted mobility and social interaction, and therefore likely to underestimate infection rates in future periods without restrictions, but is also pre-vaccination, which will overestimate infection rates. However, even if transmission peaks at one-tenth of this, it will still require a hundred-fold increase in sequencing capacity.

#### Screening in quarantine

regular testing of residents and staff linked to quarantine facilities is an important component of the surveillance system and must be continued while quarantine is in place. It gains even greater importance if the focus is early detection of imported variants, and genomic sequencing capacity cannot screen large numbers of community cases. In future, whether changes are made to quarantining arrangements or not (e.g., a shift to home quarantine etc), testing (including genomic sequencing) of all international arrivals will continue to be essential to enable early detection of VoCs. The number of international arrivals (both resident and non-resident) to Australia in late 2019, prior to COVID-19-related border closures, was over 1.5 million per month.^31^ Sequencing positive samples in this group alone would require a marked increase in capacity if positivity rates were to approximate prevalence in source countries.

#### Wastewater surveillance

Wastewater surveillance systems demonstrated limited utility over the study period in identifying and controlling novel outbreaks. Once levels of local transmission increase, there will be fewer catchments and periods where a positive sample is an unexpected finding. If the goal of surveillance shifts to identification of VoCs through sequencing of wastewater fragments,^32^ similar limitations in performance would apply. Given the limited performance in the study period, further evaluation of widespread wastewater surveillance must be done to determine its added value against other COVID-19 surveillance strategies.

### Implications for other public health control measures

Surveillance system requirements must be considered within the context of the other key components of a framework which defines the capacity to detect and control COVID-19 transmission. These are: vaccine effectiveness and uptake, outbreak response capacity (including effective implementation of social and mobility restrictions), and community engagement and partnership as the cross-cutting enabler of success in all areas. Table 2 provides a framework for future non-surveillance-related public health control measures when focus shifts to management of VoCs.

**Table 2:**
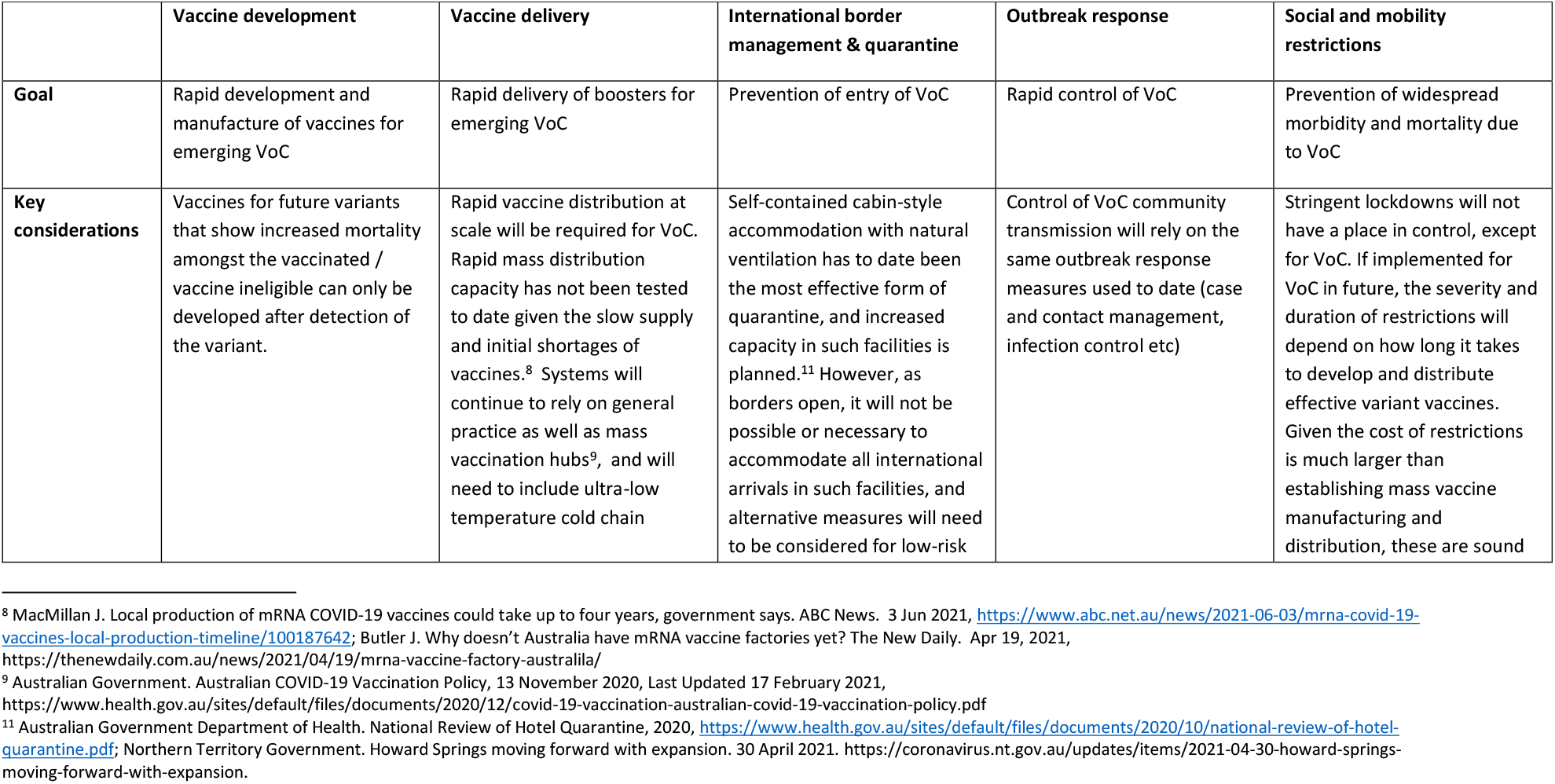
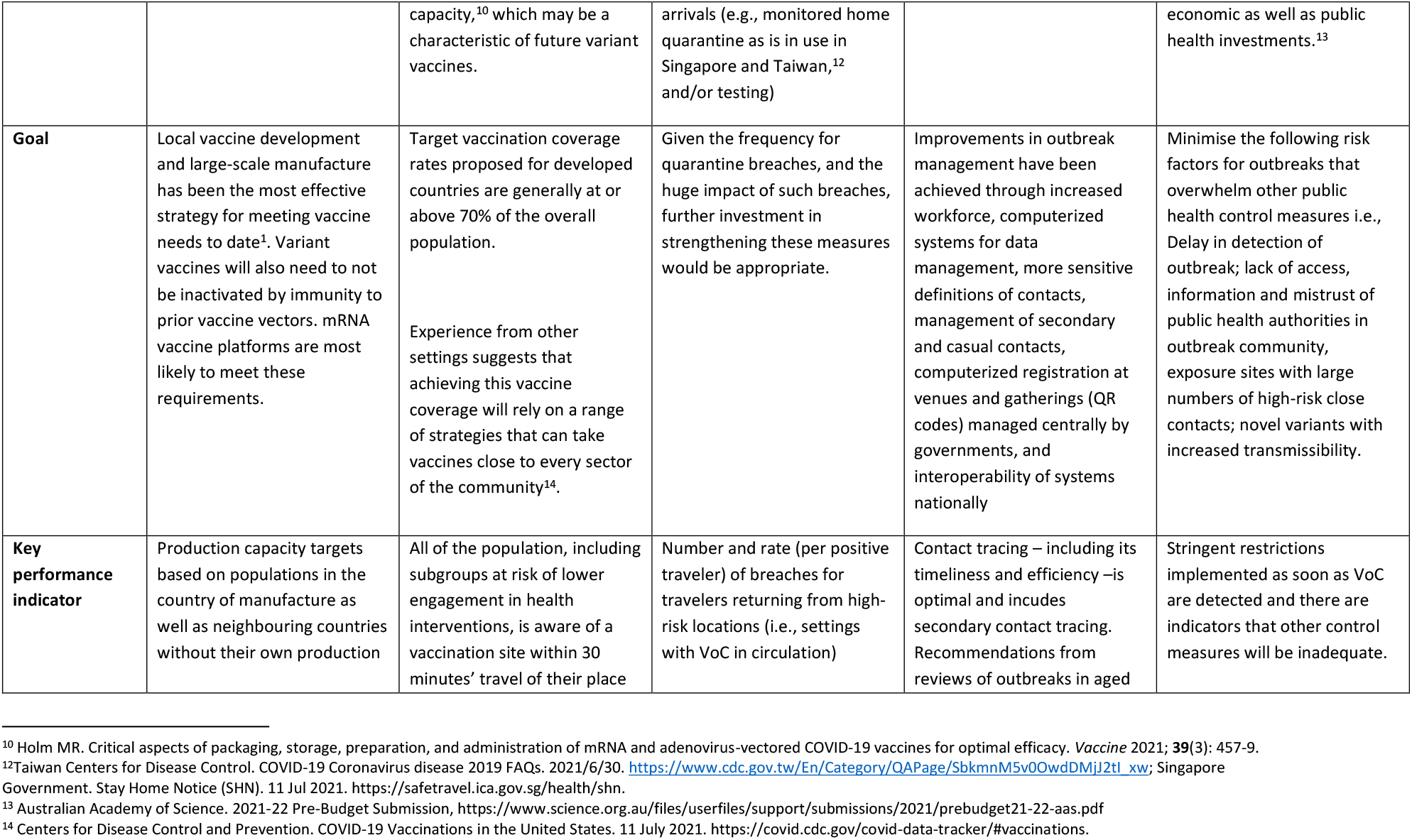
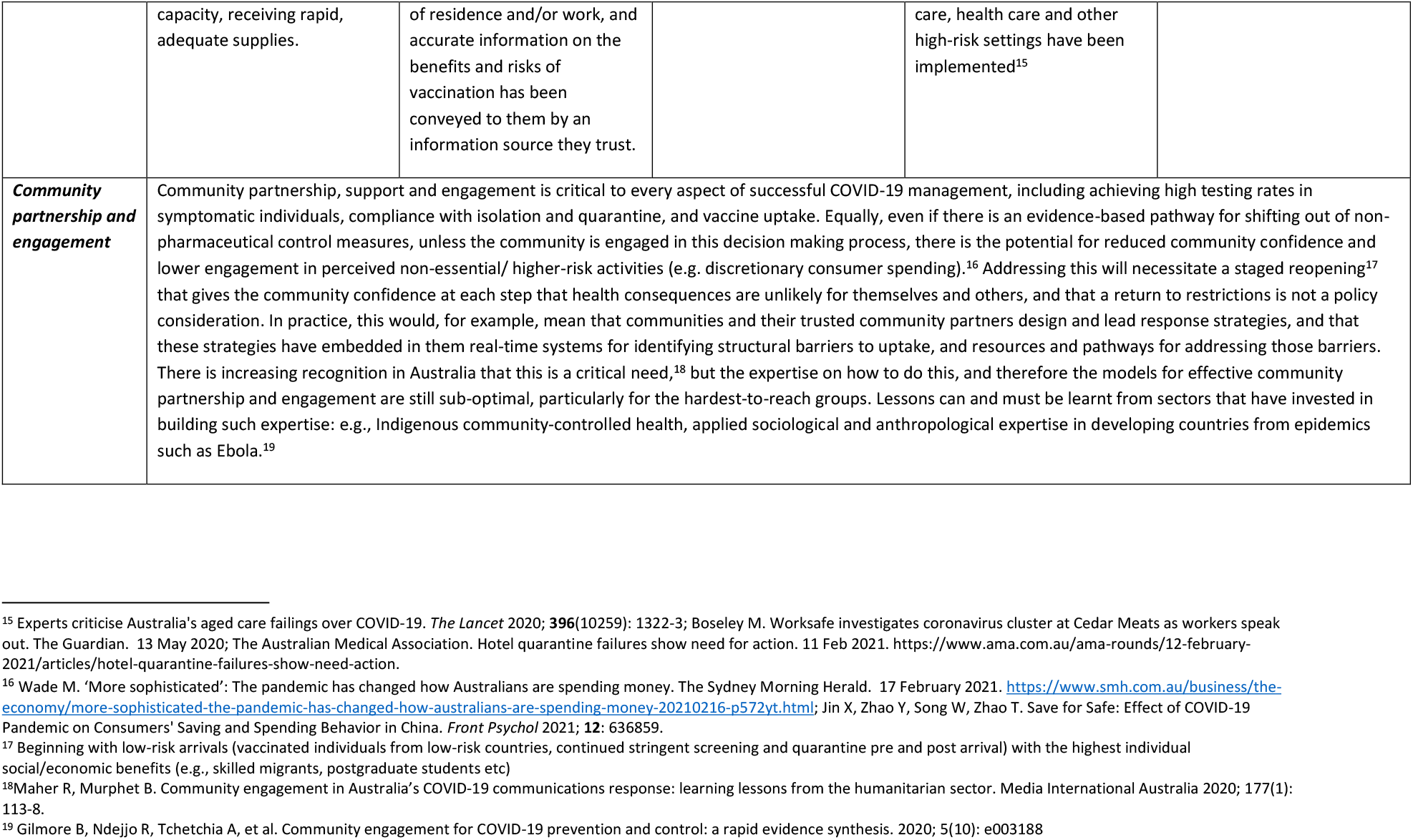
Response requirements if VoCs with increased severity, including in vaccinated and/or vaccine ineligible, were to emerge.

|  | Vaccine development | Vaccine delivery | International border management & quarantine | Outbreak response | Social and mobility restrictions |
| --- | --- | --- | --- | --- | --- |
| **Goal** | Rapid development and manufacture of vaccines for emerging VoC | Rapid delivery of boosters for emerging VoC | Prevention of entry of VoC | Rapid control of VoC | Prevention of widespread morbidity and mortality due to VoC |
| **Key considerations** | Vaccines for future variants that show increased mortality amongst the vaccinated / vaccine ineligible can only be developed after detection of the variant. | Rapid vaccine distribution at scale will be required for VoC. Rapid mass distribution capacity has not been tested to date given the slow supply and initial shortages of vaccines.<sup>8</sup> Systems will continue to rely on general practice as well as mass vaccination hubs<sup>9</sup>, and will need to include ultra-low temperature cold chain | Self-contained cabin-style accommodation with natural ventilation has to date been the most effective form of quarantine, and increased capacity in such facilities is planned.<sup>11</sup> However, as borders open, it will not be possible or necessary to accommodate all international arrivals in such facilities, and alternative measures will need to be considered for low-risk | Control of VoC community transmission will rely on the same outbreak response measures used to date (case and contact management, infection control etc) | Stringent lockdowns will not have a place in control, except for VoC. If implemented for VoC in future, the severity and duration of restrictions will depend on how long it takes to develop and distribute effective variant vaccines. Given the cost of restrictions is much larger than establishing mass vaccine manufacturing and distribution, these are sound |

<sup>8</sup> MacMillan J. Local production of mRNA COVID-19 vaccines could take up to four years, government says. ABC News. 3 Jun 2021, https://www.abc.net.au/news/2021-06-03/mrna-covid-19-vaccines-local-production-timeline/100187642; Butler J. Why doesn't Australia have mRNA vaccine factories yet? The New Daily. Apr 19, 2021, https://thenewdaily.com.au/news/2021/04/19/mrna-vaccine-factory-australila/
<sup>9</sup> Australian Government. Australian COVID-19 Vaccination Policy, 13 November 2020, Last Updated 17 February 2021, https://www.health.gov.au/sites/default/files/documents/2020/12/covid-19-vaccination-australian-covid-19-vaccination-policy.pdf
<sup>11</sup> Australian Government Department of Health. National Review of Hotel Quarantine, 2020, https://www.health.gov.au/sites/default/files/documents/2020/10/national-review-of-hotel-quarantine.pdf; Northern Territory Government. Howard Springs moving forward with expansion. 30 April 2021. https://coronavirus.nt.gov.au/updates/items/2021-04-30-howard-springs-moving-forward-with-expansion.

|  |  |  |  |  |  |
| --- | --- | --- | --- | --- | --- |
|  |  | capacity,<sup>10</sup> which may be a characteristic of future variant vaccines. | arrivals (e.g., monitored home quarantine as is in use in Singapore and Taiwan,<sup>12</sup> and/or testing) |  | economic as well as public health investments.<sup>13</sup> |
| **Goal** | Local vaccine development and large-scale manufacture has been the most effective strategy for meeting vaccine needs to date<sup>1</sup>. Variant vaccines will also need to not be inactivated by immunity to prior vaccine vectors. mRNA vaccine platforms are most likely to meet these requirements. | Target vaccination coverage rates proposed for developed countries are generally at or above 70% of the overall population.<br>Experience from other settings suggests that achieving this vaccine coverage will rely on a range of strategies that can take vaccines close to every sector of the community<sup>14</sup>. | Given the frequency for quarantine breaches, and the huge impact of such breaches, further investment in strengthening these measures would be appropriate. | Improvements in outbreak management have been achieved through increased workforce, computerized systems for data management, more sensitive definitions of contacts, management of secondary and casual contacts, computerized registration at venues and gatherings (QR codes) managed centrally by governments, and interoperability of systems nationally | Minimise the following risk factors for outbreaks that overwhelm other public health control measures i.e., Delay in detection of outbreak; lack of access, information and mistrust of public health authorities in outbreak community, exposure sites with large numbers of high-risk close contacts; novel variants with increased transmissibility. |
| **Key performance indicator** | Production capacity targets based on populations in the country of manufacture as well as neighbouring countries without their own production | All of the population, including subgroups at risk of lower engagement in health interventions, is aware of a vaccination site within 30 minutes' travel of their place | Number and rate (per positive traveler) of breaches for travelers returning from high-risk locations (i.e., settings with VoC in circulation) | Contact tracing – including its timeliness and efficiency – is optimal and includes secondary contact tracing. Recommendations from reviews of outbreaks in aged | Stringent restrictions implemented as soon as VoC are detected and there are indicators that other control measures will be inadequate. |

<sup>10</sup> Holm MR. Critical aspects of packaging, storage, preparation, and administration of mRNA and adenovirus-vectored COVID-19 vaccines for optimal efficacy. *Vaccine* 2021; **39**(3): 457-9.
<sup>12</sup>Taiwan Centers for Disease Control. COVID-19 Coronavirus disease 2019 FAQs. 2021/6/30. https://www.cdc.gov.tw/En/Category/QAPage/SbkmnM5v0OwdDMj2t1_xw; Singapore Government. Stay Home Notice (SHN). 11 Jul 2021. https://safetravel.ica.gov.sg/health/shn.
<sup>13</sup> Australian Academy of Science. 2021-22 Pre-Budget Submission, https://www.science.org.au/files/userfiles/support/submissions/2021/prebudget21-22-aas.pdf
<sup>14</sup> Centers for Disease Control and Prevention. COVID-19 Vaccinations in the United States. 11 July 2021. https://covid.cdc.gov/covid-data-tracker/#vaccinations.

|  |  |  |  |  |
| --- | --- | --- | --- | --- |
|  | capacity, receiving rapid, adequate supplies. | of residence and/or work, and accurate information on the benefits and risks of vaccination has been conveyed to them by an information source they trust. |  | care, health care and other high-risk settings have been implemented<sup>15</sup> |
| **Community partnership and engagement** | Community partnership, support and engagement is critical to every aspect of successful COVID-19 management, including achieving high testing rates in symptomatic individuals, compliance with isolation and quarantine, and vaccine uptake. Equally, even if there is an evidence-based pathway for shifting out of non-pharmaceutical control measures, unless the community is engaged in this decision making process, there is the potential for reduced community confidence and lower engagement in perceived non-essential/ higher-risk activities (e.g. discretionary consumer spending).<sup>16</sup> Addressing this will necessitate a staged reopening<sup>17</sup> that gives the community confidence at each step that health consequences are unlikely for themselves and others, and that a return to restrictions is not a policy consideration. In practice, this would, for example, mean that communities and their trusted community partners design and lead response strategies, and that these strategies have embedded in them real-time systems for identifying structural barriers to uptake, and resources and pathways for addressing those barriers. There is increasing recognition in Australia that this is a critical need,<sup>18</sup> but the expertise on how to do this, and therefore the models for effective community partnership and engagement are still sub-optimal, particularly for the hardest-to-reach groups. Lessons can and must be learnt from sectors that have invested in building such expertise: e.g., Indigenous community-controlled health, applied sociological and anthropological expertise in developing countries from epidemics such as Ebola.<sup>19</sup> |  |  |  |

<sup>15</sup> Experts criticise Australia's aged care failings over COVID-19. *The Lancet* 2020; **396**(10259): 1322-3; Boseley M. Worksafe investigates coronavirus cluster at Cedar Meats as workers speak out. *The Guardian*. 13 May 2020; The Australian Medical Association. Hotel quarantine failures show need for action. 11 Feb 2021. https://www.ama.com.au/ama-rounds/12-february-2021/articles/hotel-quarantine-failures-show-need-action.
<sup>16</sup> Wade M. 'More sophisticated': The pandemic has changed how Australians are spending money. *The Sydney Morning Herald*. 17 February 2021. https://www.smh.com.au/business/the-economy/more-sophisticated-the-pandemic-has-changed-how-australians-are-spending-money-20210216-p572yt.html; Jin X, Zhao Y, Song W, Zhao T. Save for Safe: Effect of COVID-19 Pandemic on Consumers' Saving and Spending Behavior in China. *Front Psychol* 2021; **12**: 636859.
<sup>17</sup> Beginning with low-risk arrivals (vaccinated individuals from low-risk countries, continued stringent screening and quarantine pre and post arrival) with the highest individual social/economic benefits (e.g., skilled migrants, postgraduate students etc)
<sup>18</sup> Maher R, Murphet B. Community engagement in Australia's COVID-19 communications response: learning lessons from the humanitarian sector. *Media International Australia* 2020; **177**(1): 113-8.
<sup>19</sup> Gilmore B, Ndejjo R, Tchetchia A, et al. Community engagement for COVID-19 prevention and control: a rapid evidence synthesis. 2020; **5**(10): e003188

#### Vaccination targets

A key factor defining future strategy is the expected vaccine coverage at completion of the initial vaccination program. The assumption is that, at a minimum, vaccine coverage targets must be high enough across all sections of the community, particularly in those most at risk such as the elderly, that severe disease, hospitalization, and mortality will be low and health system impacts will not be overwhelming if widespread transmission of the disease were to occur. Current Australian government policy sets a threshold of 80% of the eligible population aged 16 and over fully vaccinated as the point at which widespread, stringent social and mobility restrictions are unlikely to be required in order to maintain low levels of severe disease.^33^

However, although vaccine targets have been defined by government policy based on the percentage uptake needed across the population to achieve transmission impacts and prevent undesirable health outcomes, a more useful and ethical approach is to consider target coverage as the highest coverage achievable with available vaccines and an effective community support and engagement strategy. Such a strategy would include offering every individual in the community for whom vaccination is not contraindicated their choice of approved vaccine, in a setting and at a time that is feasible and acceptable to them, alongside material support (paid leave, free health care, including primary practitioner support) for those likely to require such support and accessible, practical, and trusted messaging and communication. The target we propose in Table 3 is based on this approach.

A key consideration is those groups at risk of lower engagement in health interventions, including vaccination, such as socially and economically disadvantaged groups. These are also the groups at higher risk of acquiring disease due to occupational and social risk factors. Although vaccine hesitancy and complacency are being referred to currently as important,^34^ past experience with vaccination suggests that the main cause for low uptake, including in these higher risk groups, is likely to be structural barriers to engagement.^35, 36^ Additionally, shifting behaviour in the small minority that are strongly vaccine resistant is likely to be complex and challenging.^37^ Again, a more useful and effective approach is that which we describe above, based on addressing structural barriers to engagement.

Another important group to consider is children. Even though younger age groups are less susceptible to severe disease, there is an ethical imperative to ensure that these groups are offered safe and effective vaccination if policies that may result in widespread transmission are adopted. Given the increased risk of severe disease now documented in children,^38^ vaccine programs are likely to include this group when eligible,^39^ with indications those 5-11 years of age will be eligible in Australia by early 2022.^40^ Also important in this regard is the impact of primary school closures and restrictions as COVID-19 transmission has moved into this unvaccinated group in the community.^41^ A final critical consideration is that any risk to children is likely to be, in the mind of the community, the most important non-negotiable in their acceptance or not of increased transmission as society opens up.

## Discussion

### Overview of main findings

Most novel incursions of SARS-CoV-2 into the Australian community have been detected through community-based surveillance for symptomatic disease, with the remainder detected through screening of returning overseas travellers and staff linked to quarantine facilities. Community-based symptomatic testing levels in the region of the outbreak were between 1-3% of the population tested in the week preceding detection of the outbreak, suggesting this level of testing will allow early detection of most community seeding or amplifying events. Outbreaks were detected, on average, within 5 days of the primary case. All outbreaks in 2021 were due to novel variants of concern. Although outbreaks due to the Delta variant of SARS-CoV-2 appear to be more transmissible and therefore more difficult to control, the surveillance system has still performed well in being able to detect them early in the course of transmission.

Wastewater surveillance was of limited utility in the study period. However in the period following the study, from July-August 2021, there were two outbreaks in which detection of the outbreak was preceded by unexpected positive wastewater surveillance results.^41-44^ These were both in regional areas without prior known cases at the time of detection, but which were adjacent to areas with extensive outbreaks. This suggests that wastewater detection may be more sensitive given variants with higher viral load such as Delta and may be of some value in areas adjacent to outbreaks that are free of transmission. Screening of wastewater from high-risk settings (e.g., public housing towers^45^) has also been utilised more recently and could continue to be utilised for early detection of novel variants from, for example, international arrivals while in the airline, and/or quarantine facility if facility-based quarantine policies are continued. However, given its overall low sensitivity and specificity, and that wastewater detection will rely on community surveillance to rapidly identify cases if it is to be of utility in public health response, investing in maintaining community surveillance through high uptake of testing will continue to be critical. Additionally, overly strong messaging focussed on unexpected wastewater detections, may, over time, result in the public presenting for testing based mainly on wastewater results, which would greatly undermine the performance of the surveillance system.

#### Genomic sequencing capacity

As our analysis suggests, to have similar performance in detecting variants as Australia has had in detecting novel outbreaks of SARS-Co-V2, sequencing rates of up to 1000-fold magnitude higher than current requirements would be required. Prioritizing sequencing to those samples linked to severe disease (hospital/ICU) would require less capacity. However previous research demonstrates that if surveillance were to rely on hospital rather than community-based case detection, the number of cases at the point of outbreak detection would be much higher, making outbreak control much more challenging.^46^ A more effective strategy would be to prioritise sequencing to international arrivals, the most likely source of novel VoC introductions in Australia. This would require ongoing comprehensive SARS-CoV-2 testing of all arrivals. Even this alone would necessitate marked scale-up of sequencing capacity if international arrival numbers were to return to pre-COVID-19 levels.

Attempts of other high-income nations to detect variants highlight current constraints on the capacity of nations to conduct large-scale genomic sequencing. As of February 2021, the US had sequenced around 96,000 (less than one percent) of its 27 million COVID-19 positive cases.^47^ Experts attribute the country’s low sequencing rate to a lack of national coordination, poor surveillance planning, and inadequate investment in sequencing infrastructure and research.^47^ In Canada, genomic surveillance has been a key component of the national surveillance strategy, with relatively greater success than the US.^48,49^ An average of five percent of COVID-19 positive samples are being analysed across the country,^50^ with a target of sequencing 10 percent of positive samples.^50^ This would bring its efforts closer to the UK, which has been lauded for its world-leading genomic sequencing capability.^51^ As of 5 July 2021, the UK has conducted approximately 672,677 sequences out of 5,186,297 infected people (13.0%) in total.^52^

However, even the UK’s level of sequencing is much lower than would be needed for Australia to conduct optimal surveillance for VoCs. Given the challenges faced by other developed countries in sequencing a high proportion of samples once the incidence of SARS-CoV-2 increases it is unlikely that full genome sequencing would be able to meet such requirements. Therefore, to implement genomic surveillance strategies that are effective in early detection of novel VoC introductions or emergence, it is critical to develop technologies that can screen for—rapidly and at scale—specific mutations of particular concern (e.g., immune escape mutations).

### Limitations

testing rates were calculated for States as a whole, and therefore may not reflect the testing rates in the specific area where the outbreak occurred. Rates vary by geographical distance from testing sites. Given all outbreaks except one were detected in major metropolitan areas, which have higher testing rates than rural areas, State-level averages are likely to be an underestimate of testing in urban areas. However, testing rates also vary widely within urban areas based on the socio-economic characteristics of the catchment population. Testing data disaggregated by LGA were not publicly available for most States, but future analysis should be done to refine these estimates if such data becomes available.

## Conclusions

Community-based surveillance for disease and routine screening of residents and staff in overseas quarantine facilities achieved early and comprehensive detection of new outbreaks of SARS-CoV-2 in the community in a period when local transmission had been eliminated in Australia and novel introductions through overseas arrivals were limited. Epidemiological investigation to identify transmission sources and link outbreak clusters has been greatly supported by comprehensive genomic sequencing.

Once vaccine coverage rates are maximised, the priority for surveillance will shift from detection of SARS-CoV-2 to emphasise detection of SARS-CoV-2 variants of concern associated with increased severity of disease in those who are vaccinated, and vaccine ineligible. This will require ongoing investment in maintaining the current community-based disease surveillance systems, as well as implementing strategies for screening of returned travellers on a larger scale. Such surveillance systems will need to be linked to greatly increased genomic sequencing capacity if novel VoCs are to be detected early in transmission. Given Australia’s limited genomic sequencing capacity, there is an urgent need for investment in scaling up and decentralizing, including through novel technologies capable of large volume throughout screening to detect mutations of concern.

Other measures to control VoCs that will need to be implemented alongside these surveillance strategies include maintaining and improving the efficiency of outbreak response, and developing the capacity to rapidly engineer, manufacture, and distribute variant vaccines at scale. The most important factor in prevention, detection, and control of current and future transmission, including our capacity to open society safely, will be how well governments engage with and support all sectors of the community. Investment in applied research on the structural barriers to engagement, and how these need to be addressed, is critical to ensuring success against COVID-19. Finally, all the measures we have discussed are aimed at mitigating novel variants of concern, the main risk to future control of COVID-19 in highly vaccinated populations. However, the only way to prevent their emergence is to support high vaccine coverage globally.

## Supporting information

STARI Checklist

COI Disclosure Street

COI Disclosure Banks

COI Disclosure DOnise

COI Disclosure Glass

COI Disclosure Jantos

COI Disclosure Lokuge

COI Disclosure Baptista

## Data Availability

All data produced in the present study are available upon reasonable request to the authors

## Ethics

Ethical approval was not required for this study as it involved no human or animal participation. The research was conducted according to the Australian Code for the Responsible Conduct of Research.

## References

1. Bremmer I. The Best Global Responses to the COVID-19 Pandemic, 1 Year Later: Time; [updated 23 Feb 2021. Available from: https://time.com/5851633/best-global-responses-covid-19/.

2. Lowy Institute. Covid Performance Index 2021 [Available from: https://interactives.lowyinstitute.org/features/covid-performance/#overview.

3. Communicable Diseases Network Australia. Australian National Disease Surveillance Plan for COVID-19, Version 2.0, April 2021. 2021.

4. Australian Government Department of Health, Communicable Diseases Network Australia. Coronavirus Disease 2019 (COVID-19) - CDNA National Guidelines for Public Health Units, Version 4.7. 2021.

5. Hong J, Chang R, Varley K. The best and worst places to be in the Coronavirus era: Bloomberg; 2020 [updated 25 May 2021. Available from: https://www.bloomberg.com/graphics/covid-resilience-ranking/.Google

6. Grout L, Katar A, Ouakrim DA, Summers JA, Kvalsvig A, Baker MG, Blakely T, et al. Estimating the Failure Risk of Quarantine Systems for Preventing COVID-19 Outbreaks in Australia and New Zealand. 2021.

7. Duckett S, Stobart A. Australia’s COVID-19 response: the four successes and four failures: Grattan Institute; 2020 [updated 12 June 2020. Available from: https://grattan.edu.au/news/australias-covid-19-response-the-four-successes-and-four-failures/.

8. Abdool Karim SS, de Oliveira T. New SARS-CoV-2 Variants — Clinical, Public Health, and Vaccine Implications. New England Journal of Medicine. 2021;384(19):1866–8.

9. World Health Organization. COVID-19 Vaccines 2021 [Available from: https://www.who.int/emergencies/diseases/novel-coronavirus-2019/covid-19-vaccines.

10. World Health Organization. Coronavirus disease (COVID-19): Vaccines 2020 [Available from: https://www.who.int/news-room/q-a-detail/coronavirus-disease-(covid-19)-vaccines.

11. Public Health England. Public Health England vaccine effectiveness report. 2021 March 2021.

12. Time. COVID-19 Vaccine Tracker [updated 08/07/2021. Available from: https://time.com/collection/coronavirus-vaccines-updates/.

13. The Economist. More than 85 poor countries will not have widespread access to coronavirus vaccines before 2023 [updated 27 Jan 2021. Available from: https://www.eiu.com/n/85-poor-countries-will-not-have-access-to-coronavirus-vaccines/.

14. World Health Organization. WHO Director-General’s opening remarks at 148th session of the Executive Board [updated 18 January 2021. Available from: https://www.who.int/director-general/speeches/detail/who-director-general-s-opening-remarks-at-148th-session-of-the-executive-board.

15. Burki T. Global COVID-19 vaccine inequity. The Lancet Infectious Diseases. 2021;21(7):922–3.

16. World Health Organization. COVID-19 Weekly Epidemiological Update. 6 July 2021.

17. World Health Organization. Report of the WHO-China Joint Mission on Coronavirus Disease 2019 (COVID-19). 16–24 February 2020.

18. Grant MC, Geoghegan L, Arbyn M, Mohammed Z, McGuinness L, Clarke EL, Wade RG. The prevalence of symptoms in 24,410 adults infected by the novel coronavirus (SARS-CoV-2; COVID-19): A systematic review and meta-analysis of 148 studies from 9 countries. PLoS One. 2020;15(6):e0234765.

19. Zwartz H. A lot more of us can now get tested for coronavirus. Here’s what you need to know. ABC News. 24 Apr 2020.

20. Grundy E, Slattery P, Buckland G, Suresh Babu S, Mangiarulo M, Oren P, Dillon C. Scrub Project Wave 4: Australians’ Views on Private Gatherings, Remote Working and Getting Tested: Monash University; [updated 24 June 2020. Available from: https://www.behaviourworksaustralia.org/scrub-project-wave-4-australians-views-on-private-gatherings-remote-working-and-getting-tested/.

21. Andersson P, Sherry NL, Howden BP. Surveillance for SARS-CoV-2 variants of concern in the Australian context. Medical Journal of Australia. 2021.

22. Australian Government Department of Health. Coronavirus (COVID-19) advice for international travellers [updated 7 July 2021. Available from: https://www.health.gov.au/news/health-alerts/novel-coronavirus-2019-ncov-health-alert/coronavirus-covid-19-restrictions/coronavirus-covid-19-advice-for-international-travellers#quarantine-for-incoming-travellers.

23. Rules on COVID vaccines, testing for quarantine workers tightened after National Cabinet meeting. ABC News. 29 Jun 2021.

24. Associated Press. Israel to offer Pfizer Covid booster shots to people over 60: The Guardian; [updated 30 July 2021. Available from: https://www.theguardian.com/world/2021/jul/30/israel-to-offer-pfizer-covid-booster-shots-to-people-over-60.

25. World Health Organization. COVID-19 Weekly Epidemiological Update. 2021 22 June 2021.

26. The Agency for Clinical Innovation, NSW Government. COVID-19 Critical Intellgence Unit: Living Evidence - SARS-CoV-2 variants [updated 12 Jul 2021. Available from: https://aci.health.nsw.gov.au/covid-19/critical-intelligence-unit/sars-cov-2-variants.

27. Torjesen I. Covid-19: Delta variant is now UK’s most dominant strain and spreading through schools. BMJ. 2021;373:n1445.

28. Paddock R, Suhartono M. No Longer ‘Hidden Victims,’ Children Are Dying as Virus Surges in Indonesia: New York Times; [updated 31 July 2021. Available from: https://www.nytimes.com/2021/07/25/world/asia/children-deaths-virus-indonesia.html.

29. Gottlieb RL, Nirula A, Chen P, Boscia J, Heller B, Morris J, Huhn G, et al. Effect of Bamlanivimab as Monotherapy or in Combination With Etesevimab on Viral Load in Patients With Mild to Moderate COVID-19. JAMA. 2021;325(7):632.

30. Centers for Disease Control and Prevention. Estimated COVID-19 Burden. US Department of Health and Human Services; 2021. Table 2 from CDC: Estimated rates of COVID-19 disease outcomes per 100,000, by age group — United States, February 2020-May 2021 [updated 26 Aug 2021, cited 27 Aug 2021]. Available from: https://www.cdc.gov/coronavirus/2019-ncov/cases-updates/burden.html

31. Australian Bureau of Statistics. Overseas Arrivals and Departures, Australia. Australian Bureau of Statistics; 16 Jan 2020. Available from: https://www.abs.gov.au/statistics/industry/tourism-and-transport/overseas-arrivals-and-departures-australia/nov-2019.

32. Bar-Or I, Weil M, Indenbaum V, Bucris E, Bar-Ilan D, Elul M, Levi N, et al. Detection of SARS-CoV-2 variants by genomic analysis of wastewater samples in Israel. Science of The Total Environment. 2021;789:148002.

33. Morrison S. Press Conference - Canberra, ACT, Prime Minister (Transcript) [updated 30 Jul 2021. Available from: https://www.pm.gov.au/media/press-conference-canberra-act-9.

34. Tingle L. If the public has vaccine hesitancy, the government has developed strategy hesitancy: ABC News; [updated 22 May 2021. Available from: https://www.abc.net.au/news/2021-05-22/if-public-has-vaccine-hesitancy-government-strategy-hesitancy/100154798.

35. Anderson EL. Recommended solutions to the barriers to immunization in children and adults. Mo Med. 2014;111(4):344–8.

36. Leask J. Being open about why Australia’s vaccination take-up is low is the first step to improve it: The Guardian; [updated 25 May 2021. Available from: https://www.theguardian.com/commentisfree/2021/may/25/being-open-about-why-australias-vaccination-take-up-is-low-is-the-first-step-to-improve-it.

37. Murphy J, Vallières F, Bentall RP, Shevlin M, McBride O, Hartman TK, McKay R, et al. Psychological characteristics associated with COVID-19 vaccine hesitancy and resistance in Ireland and the United Kingdom. Nature Communications. 2021;12(1):29.

38. Havers F, Whitaker M, Self J, et al. Hospitalization of Adolescents Aged 12–17 Years with Laboratory-Confirmed COVID-19 — COVID-NET, 14 States, March 1, 2020–April 24, 2021. MMWR Morb Mortal Wkly Rep. 2021;70(23):851–7.

39. Lovelace BJ. Moderna says it plans to expand Covid vaccine trial for kids 5 to 11, will seek FDA OK as early as year-end: CNBC; [updated 26 Jul 2021. Available from: https://www.cnbc.com/2021/07/26/covid-vaccine-moderna-says-it-plans-to-expand-trial-for-kids-5-to-11.html.

40. Ministers, Department of Health [radio transcript]. Department of Health; 2021. Interview with Jim Wilson on 2GB Drive, on 12 November 2021, on Australia’s vaccination rollout. 13 Nov 2021. Available from: https://www.health.gov.au/ministers/the-hon-greg-hunt-mp/media/interview-with-jim-wilson-on-2gb-drive-on-12-november-2021-on-australias-vaccination-rollout.

41. New South Wales Department of Health. COVID-19 (Coronavirus) statistics 04 August 2021 [Available from: https://www.health.nsw.gov.au/news/Pages/20210804_01.aspx.

42. New South Wales Department of Health. COVID-19 (Coronavirus) statistics 05 August 2021 [Available from: https://www.health.nsw.gov.au/news/Pages/20210805_00.aspx.

43. Queensland Government. Wastewater surveillance program results 09 August 2021 [Available from: https://www.qld.gov.au/health/conditions/health-alerts/coronavirus-covid-19/current-status/wastewater.

44. Queensland Government. Update – COVID-19 08 August 2021 [Available from: https://www.health.qld.gov.au/news-events/doh-media-releases/releases/update-covid-23.

45. Victoria State Government. Coronavirus update for Victoria – 19 August 2021. Available from: https://www.dhhs.vic.gov.au/coronavirus-update-victoria-19-august-2021.

46. Lokuge K, Banks E, Davis S, Roberts L, Street T, O’Donovan D, Caleo G, et al. Exit strategies: optimising feasible surveillance for detection, elimination, and ongoing prevention of COVID-19 community transmission. BMC Medicine. 2021;19(1):50.

47. Thulin L. Why the U.S. Is Struggling to Track Coronavirus Variants. Smithsonian Magazine. 11/02/2021.

48. Government of Canada invests $53 million to address COVID-19 virus variants of concern [press release]. https://www.canada.ca/en/public-health/news/2021/02/government-of-canada-invests-53-million-to-address-covid-19-virus-variants-of-concern.html: Government of Canada, 12/02/2021 2021.

49. Genome Canada. CanCOGeN 2021 [Available from: https://www.genomecanada.ca/en/cancogen.

50. Bains C. Canada’s hunt for COVID-19 variants to ramp up with more genomic sequencing: Global News; [updated 19/02/2021. Available from: https://globalnews.ca/news/7650061/canada-covid-genomic-sequencing-variants/.

51. UK Department of Health and Social Care. UK exceeds 600,000 COVID-19 tests genomically sequenced (press release) [updated 2 July 2021. Available from: https://www.gov.uk/government/news/uk-exceeds-600000-covid-19-tests-genomically-sequenced.

52. COVID-19 Genomics UK Consortium. Summary report: COG-UK geographic coverage of SARS-CoV-2 sample sequencing. 5 July 2021.

