## Supplementary material for "Opening up safely: public health system requirements for ongoing COVID-19 management based on evaluation of Australia’s surveillance system performance": STARI Checklist

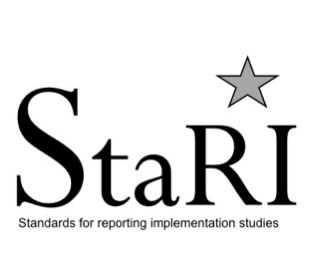
**Standards for Reporting Implementation Studies: the StaRI checklist for completion**

The StaRI standard should be referenced as: Pinnock H, Barwick M, Carpenter C, Eldridge S, Grandes G, Griffiths CJ, Rycroft-Malone J, Meissner P, Murray E, Patel A, Sheikh A, Taylor SJC for the StaRI Group. Standards for Reporting Implementation Studies [(StaRI) statement](http://www.bmj.com/content/356/bmj.i6795.full). *BMJ* 2017;356:i6795

The detailed Explanation and Elaboration document, which provides the rationale and exemplar text for all these items is: Pinnock H, Barwick M, Carpenter C, Eldridge S, Grandes G, Griffiths C, Rycroft-Malone J, Meissner P, Murray E, Patel A, Sheikh A, Taylor S, for the StaRI group. Standards for Reporting Implementation Studies [(StaRI). Explanation and Elaboration document](http://bmjopen.bmj.com/content/7/4/e013318.full?ijkey=vv4LKZxc25YcLJv&keytype=ref). *BMJ Open* 2017 2017;7:e013318

Notes: A key concept of the StaRI standards is the dual strands of describing, on the one hand, the implementation strategy and, on the other, the clinical, healthcare, or public health intervention that is being implemented. These strands are represented as two columns in the checklist.

| The primary focus of implementation science is the implementation strategy (column 1) and the expectation is that this will always be completed. | The evidence about the impact of the intervention on the targeted population should always be considered (column 2) and either health outcomes reported or robust evidence cited to support a known beneficial effect of the intervention on the health of individuals or populations. |
| --- | --- |

The StaRI standardsrefers to the broad range of study designs employed in implementation science. Authors should refer to other reporting standards for advice on reporting specific methodological features. Conversely, whilst all items are worthy of consideration, not all items will be applicable to, or feasible within every study.

| **Checklist item** | | **Reported on page #** | **Implementation Strategy** | **Reported on page #** | **Intervention** |
| --- | --- | --- | --- | --- | --- |
|  | |  | “Implementation strategy” refers to how the intervention was implemented |  | “Intervention” refers to the healthcare or public health intervention that is being implemented. |
| **Title and abstract** | | | | | |
| Title | **1** | 1 | Identification as an implementation study, and description of the methodology in the title and/or keywords | | |
| Abstract | **2** | 1-2 | Identification as an implementation study, including a description of the implementation strategy to be tested, the evidence-based intervention being implemented, and defining the key implementation and health outcomes. | | |
| **Introduction** | | | | | |
| Introduction | **3** | 4 | Description of the problem, challenge or deficiency in healthcare or public health that the intervention being implemented aims to address. | | |
| Rationale | **4** | 3-4 | The scientific background and rationale for the implementation strategy (including any underpinning theory/framework/model, how it is expected to achieve its effects and any pilot work). | 3-4, 6-7 | The scientific background and rationale for the intervention being implemented (including evidence about its effectiveness and how it is expected to achieve its effects). |
| Aims and objectives | **5** | 4 | The aims of the study, differentiating between implementation objectives and any intervention objectives. | | |
| **Methods: description** | | | | | |
| Design | **6** | 5-6 | The design and key features of the evaluation, (cross referencing to any appropriate methodology reporting standards) and any changes to study protocol, with reasons | | |
| Context | **7** | 4 | The context in which the intervention was implemented. (Consider social, economic, policy, healthcare, organisational barriers and facilitators that might influence implementation elsewhere). | | |
| Targeted ‘sites’ | **8** |  | The characteristics of the targeted ‘site(s)’ (e.g locations/personnel/resources etc.) for implementation and any eligibility criteria. |  | The population targeted by the intervention and any eligibility criteria. |
| Description | **9** | 5 | A description of the implementation strategy | 6-7 | A description of the intervention |
| Sub-groups | **10** |  | Any sub-groups recruited for additional research tasks, and/or nested studies are described | | |
| **Methods: evaluation** | | | | | |
| Outcomes | **11** | 4 | Defined pre-specified primary and other outcome(s) of the implementation strategy, and how they were assessed. Document any pre-determined targets | 6 | Defined pre-specified primary and other outcome(s) of the intervention (if assessed), and how they were assessed. Document any pre-determined targets |
| Process evaluation | **12** | 4 | Process evaluation objectives and outcomes related to the mechanism by which the strategy is expected to work | | |
| Economic evaluation | **13** |  | Methods for resource use, costs, economic outcomes and analysis for the implementation strategy |  | Methods for resource use, costs, economic outcomes and analysis for the intervention |
| Sample size | **14** |  | Rationale for sample sizes (including sample size calculations, budgetary constraints, practical considerations, data saturation, as appropriate) | | |
| Analysis | **15** | 4-6 | Methods of analysis (with reasons for that choice) | | |
| Sub-group analyses | **16** |  | Any a priori sub-group analyses (e.g. between different sites in a multicentre study, different clinical or demographic populations), and sub-groups recruited to specific nested research tasks | | |
| **Results** | | | | | |
| Characteristics | **17** | 6-7 | Proportion recruited and characteristics of the recipient population for the implementation strategy | 6-7 | Proportion recruited and characteristics (if appropriate) of the recipient population for the intervention |
| Outcomes | **18** | 7-9 | Primary and other outcome(s) of the implementation strategy | 9-10 | Primary and other outcome(s) of the Intervention (if assessed) |
| Process outcomes | **19** | 10-13 | Process data related to the implementation strategy mapped to the mechanism by which the strategy is expected to work | | |
| Economic evaluation | **20** |  | Resource use, costs, economic outcomes and analysis for the implementation strategy |  | Resource use, costs, economic outcomes and analysis for the intervention |
| Sub-group analyses | **21** |  | Representativeness and outcomes of subgroups including those recruited to specific research tasks | | |
| Fidelity/ adaptation | **22** |  | Fidelity to implementation strategy as planned and adaptation to suit context and preferences |  | Fidelity to delivering the core components of intervention (where measured) |
| Contextual changes | **23** |  | Contextual changes (if any) which may have affected outcomes | | |
| Harms | **24** |  | All important harms or unintended effects in each group | | |
| **Discussion** | | | | | |
| Structured discussion | **25** | 13 | Summary of findings, strengths and limitations, comparisons with other studies, conclusions and implications | | |
| Implications | **26** | 13-15 | Discussion of policy, practice and/or research implications of the implementation strategy (specifically including scalability) | 13-15 | Discussion of policy, practice and/or research implications of the intervention (specifically including sustainability) |
| **General** | | | | | |
| Statements | **27** | 16 | Include statement(s) on regulatory approvals (including, as appropriate, ethical approval, confidential use of routine data, governance approval), trial/study registration (availability of protocol), funding and conflicts of interest | | |
