## Supplementary Material for "Opening up safely: public health system requirements for ongoing COVID-19 management based on evaluation of Australia’s surveillance system performance"

### Supplementary Materials

##### 1. Definitions

***Testing rates*** were defined as the proportion of the total population in that State tested for Sars-CoV-2. These were calculated for the week preceding the detection of the outbreak, for the State in which the outbreak occurred.

***Outbreak definition:*** Outbreaks of COVID-19 included in this analysis included any in which one or more cases were due to community (locally-acquired) transmission within the specified time period from 1^st^ November 2020 to 30^th^ June 2021. Community transmission included any local transmission in the community, as well as any cases in quarantine or health workers in direct contact with international travellers. It also included international travellers who were infected while in quarantine in Australia, but excluded those international travellers who were infected prior to entering quarantine or who were infected by household/family contacts with whom they were residing while in quarantine. Each individual outbreak was defined as all cases epidemiologically linked to each other. If epidemiological links were not identified between distinct clusters of cases, these clusters were still considered different outbreaks even if a common source in a returned overseas traveller was identified through genomic sequencing.

***Index case:*** The index case was defined as the first reported case identified in that outbreak.

***Source case:*** The source case was defined as the case with the earliest date of symptom onset and/or infectivity that could be linked to that outbreak through genomic evidence.

***Primary case:*** The primary case was defined as the case with the earliest date of symptom onset and/or infectivity that could be epidemiologically linked to that outbreak.

(Note: in general, the epidemiological terms ‘primary’ and ‘source’ case are used interchangeably as they are the same individual. However, in this paper we have differentiated these 2 terms and adopted slightly different definitions, as described above, in order to account for the fact that the true primary /source case, which was always an international arrival) was often not linked epidemiologically, despite being linked through genomics. By differentiating, we were able to better describe the characteristics of the outbreaks and their detection).

##### 2. Characteristics of community SARS-CoV-2 outbreaks, **Australia, November 2020-June 2021**

| **Location, dates^*^** | **Cases** | **Outbreak or cluster** | **Epi link source and primary** | **Date first notified** | **Date primary case infectious** | **Source** | **First identified** | **Identified through community or quarantine** | **Genomic variant** | **Restrictions at time of detection** | **State testing rate in preceding week** |
| --- | --- | --- | --- | --- | --- | --- | --- | --- | --- | --- | --- |
| SA (Adelaide) 8/11/2020-23/12/2020 | 34^1^ | O | No epi link^3^ | 15/11/2020^1^ | 08/11/2020^3^ | Overseas traveller who infected quarantine worker^3^ | Symptomatic parent of quarantine worker tested on presentation to hospital^3^ | Community^3^ | Not reported | No major restrictions^1^ | 0.920%^2^ |
| NSW (Northern Beaches) 10/12/2020-14/01/2021 | Approximately 151^8^ | O | No epi link | 16/12/2020^7^ | Not reported | Overseas traveller who arrived 01/12/2020^7^ | Symptomatic elderly couple in community 10/12/2020^7^ | Community^7^ | Reported as most likely a strain from the US^7^ | No major restrictions^6^ | 0.892%^4,5^ |
| NSW (Quarantine) 22/12/2020-16/01/2021 | 4^9^ | O | Epi link | 22/12/2020^10^ | 14/12/2020 | Overseas traveller^12^ | Patient transport worker transporting patients from Sydney Airport to hotel quarantine ^11,12^ | Quarantine^12^ | Not reported | No major restrictions^10^ | 1.033%^2^ |
| NSW (Croydon) 28/12/2020-14/01/2021 | 11^14^ | O | No epi link | 28/12/2020^13^ | Not reported | Northern Beaches cluster^13^ | 3 adults and 3 children in one family across multiple households tested positive^13^ | Community^13^ | Genomically linked to the Northern Beaches cluster^16^ | Limited access to Northern Beaches area^15^ | 3.354%^2,5^ |
| VIC (Melbourne, Blackrock) 30/12/2020-21/01/2021 | 27^18^ | O | No epi link | 30/12/2020^17,18^ | 19/12/202019,^18^ | Thought to be an overseas traveller; genomically linked to NSW clusters; Index case had NSW, QLD travel^17,18^ | Close/casual contacts of returned traveller linked to Melbourne restaurant^17,18^ | Community^17,18^ | Not reported | Not reported | 0.941%^2^ |
| NSW (Berala) 31/12/2020-15/01/2021 | 28 | C | Epi link | 31/12/2020^19^ | 21/12/2020^19^ | The patient transport worker listed in outbreak ‘NSW Quarantine’ above^19,20^ | Berala bottle shop employee who served a colleague of the source case, working several shifts afterward^20,21^ | Community^20^ | Not reported | Limited access to Northern Beaches area^15^ | 3.044%^2^ |
| VIC (Melbourne, Vermont South) 05/01/2021 – 19/01/2021 | 1^22^ | O | No epi link | 05/01/2021^22^ | Not reported | Unknown - genomic testing shows connection to NSW strain from Sydney’s Northern Beaches outbreak^22,23^ | Symptomatic VIC resident tested positive despite no NSW travel or attendance at high-risk VIC exposure sites^22,23^ | Community^22^ | Not reported | Not reported | - |
| QLD (Brisbane quarantine hotel) 07/01/2021 – 27/01/2021 | 6^28^ | O | Epi link | 07/01/2021^27^ | 02/01/2021^25^ | Overseas traveller at the Grand Chancellor Hotel^25,27^ | Infectious quarantine worker identified in weekly testing of quarantine staff^25,27,29^ | Quarantine^25,29^ | Alpha variant^27^ | No major restrictions^26,28^ | 3.271%^2,24^ |
| VIC (Melbourne, Park Royal)  24/01/2021 – 07/02/2021 | 6^30^ | O | Epi link | 24/01/2021^30^ | Not reported | Returned overseas traveller^30^ | Family of 5 travellers test positive, source case is traveller in opposite room^30^ | Quarantine^30^ | Alpha variant (Variant of Concern)^30^ | Not reported | 1.686%^2^ |
| VIC (Melbourne, Grand Hyatt) 03/02/2021 - 17/02/2021 | 1^31^ | O | Epi link | 03/02/2021^32,33^ | 29/01/2021^32^ | Returned overseas traveller^32^ | 03/02/2021 Quarantine worker tested positive 4 days after onset, 3 days after last test^31,32,33^ | Community^33^ | Alpha variant (Variant of Concern)^34^ | Not reported | 1.304%^2^ |
| VIC (Melbourne, Holiday Inn) 07/02/2021 – 14/03/2021^39^ | 11^35,36,37^ | O | Epi link | 07/02/2021^35,37^ | Not reported | Returned overseas traveller^35,37^ | Quarantine worker tested positive during routine testing^35,37^ | Quarantine^35,37^ | Variant of Concern^37^ | Not reported | 1.342%^2^ |
| VIC (Melbourne, Coburg) 10/02/2021-14/03/2021 | 13^39,40^ | C | Epi link | 10/02/202^39,40^ | Not reported | Overseas traveller with a case of a Variant of Concern in Holiday Inn outbreak^40^ | Quarantine worker who’d visited the Holiday Inn (outbreak above) tested negative on 3, 4 and 7 Feb, then transmitted at gathering on 6 Feb and tested positive on 10/02/2021^39,40^ | Community^40^ | Alpha variant (Variant of Concern)^38^ | Not reported | 1.831%^2^ |
| QLD (Brisbane Princess Alexandra Hospital) 12/03/2021-29/03/2021 | 3 | O | Epi link | 12/03/2021^42^ | 11/03/2021^42^ | Overseas traveller^42^ | Quarantine-related healthcare worker who treated source case^42^ | Community^42^ | Alpha variant (Variant of Concern)^42^ | No major restrictions^41^ | 0.826%^2,24^ |
| QLD (Brisbane, Princess Alexandra Hospital) Related to previous cluster 29/03/2021 - 15/04/2021 | 13^46^ | C | Epi link | 29/03/2021^47^ | 23/03/2021^45^ | Overseas traveller who was a patient at the hospital tested positive on 22/03/2021^43^ | Healthcare worker at the hospital - likely to have been infected on 23/03/2021^43^ | Community^44^ | Alpha variant (Variant of Concern)^43^ | No major restrictions^45,47^ | 1.281%^2^ |
| WA  (Perth and Peel) 23/04/2021 - 15/05/2021 | 3^48^ | O | Epi link | 23/04/2021^48^ | 17/04/2021^48^ | Overseas traveller infected during quarantine^48^ | Overseas traveller infectious in community for 5 days including a flight to Melbourne^48^ | Community^48^ | Not reported | No major restrictions^49^ | 0.865%^2^ |
| WA  (Perth) 27/04/2021 – 22/05/2021 | 3^50,53^ | O | Epi link | 01/05/2021^50,52^ | 27/04/2021^50,52^ | Overseas traveller at the Pan Pacific Quarantine Hotel^52^ | Quarantine hotel security guard who worked during transfer of source case to quarantine^52^ | Quarantine^52^ | Not reported | No major restrictions^51^ | 2.582%^2^ |
| NSW  (Eastern Sydney) 05/05/2021 - 19/05/2021 | 2^54,56^ | O | No epi link | 05/05/2021^55^ | Not reported | Overseas traveller transferred from the Park Royal Hotel on 28/04/2021 ^56^ | Resident of eastern Sydney^55^ | Community^55^ | Delta variant^56^ | No major restrictions^57^ | 0.926%^2^ |
| VIC (Melbourne, Whittlesea) 23/05/2021 – 21/06/2021 | 32^60^ | O | No epi link | 23/05/2021^58^ | 20/05/2021^62^ | Genomic link to South Australian medihotel outbreak and positive case who tested positive on 10/06/2021, symptomatic as of 08/06/2021 in Wollert^59,62^ | 2 individuals in Whittlesea who tested positive on 23/05/2021; one was symptomatic as of Thursday 20/05/2021, the other was asymptomatic; genomically connected to source case but no evidence on epi link^58,62^ | Community^58^ | Kappa variant^61^ | No major restrictions^63^ | 1.814%^2^ |
| VIC  (Port Melbourne cluster) 26/05/2021 – 23/06/2021 | 32^60^ | C | No epi link | 26/05/2021^65^ | Not reported | Individual from the Whittlesea outbreak (genomically linked to SA medihotel outbreak/positive Wollert case) who worked in Port Melbourne^59,62,64^ | Co-worker of source case who also visited their Port Melbourne workplace and was considered a primary close contact^64^ | Community | Kappa variant^61^ | Not reported | 1.782%^2^ |
| VIC  (Arcare Maidstone aged care facility) 28/05/2021 – 03/07/2021 (ongoing as at 30/06/2021) | 13^66^ | O | No epi link | 30/05/2021^67^ | 26/05/2021^67^ | Genomic link to SA medi hotel outbreak and positive case in Wollert^67^ | Employee at the aged care centre^67^ | Community^67^ | Kappa variant^61^ | Lockdown with 5km radius as of 27/05/2021^68^ | 3.163%^2^ |
| VIC  (West Melbourne cluster) 01/06/21 –22/06/2021 | 15^60^ | O | No epi link | 01/06/2021^70^ | Could have been as early as 23/05/2021^71^ | Overseas traveller in quarantine who tested positive on 08/05/2021^70,71^ | Family of 4 from West Melbourne who travelled to Jervis Bay from 19-24/05/2021^70^ | Community^70^ | Delta variant^69,71^ | Lockdown with 5km radius as of 27/05/2021^68^ | 4.111%^2^ |
| VIC (Reservoir household) 09/06/2021 – 28/06/2021 | 4^73^ | O | No epi link | 09/06/2021^73^ | Not reported | Unknown; genomic link to Whittlesea outbreak^72^ | Household resident in reservoir^73^ | Community^73^ | Kappa variant^72^ | Lockdown with eased restrictions including 25km radius^74^ | 3.845%^2^ |
| VIC (Interstate travellers) 09/06/2021 – 24/06/2021 | 2^75,76^ | O | No epi link | 09/06/2021^75,76^ | Not reported | Unknown; genomic link to Whittlesea outbreak^72^ | Individual travelling with partner from Melbourne to the Sunshine Coast^75^ | Community^75^ | Kappa variant ^72^ | Lockdown with eased restrictions including 25km radius^74^ | 3.722%^2^ |
| VIC  (Kings Park, Southbank) 11/06/2021 – 10/07/2021 (ongoing as at 30/06/2021) | 12^78^ | O | Epi link | 11/06/2021^77,79^ | Not reported | A case from Arcare Aged Care Facility outbreak living in the Kings Park apartment complex transmitted virus to another resident^77,80^ | Another resident of Kings Park complex^77,79^ | Community^79^ | Not reported | Lockdown with eased restrictions including 25km radius^74^ | 3.216%^2^ |
| NSW  (Eastern Suburbs - June cluster) 16/06/2021 - ongoing | 15 as at 22/06/2021^81^ | O | No epi link | 16/06/2021^82,87^ | In the fortnight leading up to 11/06/2021^83^ | Unknown as at 24/06/2021, may have been flight crew member from the US^82^; there is no evidence of a link between this outbreak and previous NSW eastern suburbs outbreak | Driver transporting international flight crews tested positive on 16/06/2021 but undertook their first saliva swab on 15/06, which was found later to be positive^82,87^ | Quarantine (transport)^82^ | Delta B.1.617.2 variant^82,85^ | No major restrictions^86^ | 1.448%^2,84^ |
| QLD  (Portuguese Family Centre Cluster) 19/06/2021 – 21/07/2021 (ongoing as at 30/06/2021) | 7 cases as at 25/06/2021^92^ | O | Epi link | 19/06/2021^88^ | 19/06/2021^88^ | Overseas traveller tested positive a few hours after leaving 14-day hotel quarantine^88^ | The source case who had entered the community^88^ | Quarantine^91^ | Alpha variant^89^ | No major restrictions^90^ | 1.075%^2,24^ |
| NT  (Central Australian mine cluster)  25/06/2021 – 12/07/2021 | 7 as at 28/06/2021^88^ | O | Epi link | 25/06/2021^97^ | 18/06/2021^95,97^ | Interstate FIFO miner who travelled from regional VIC to Brisbane where they stayed in a quarantine hotel and contracted virus on 18/06/2021, and then worked at the mine while infectious^95,97^ | Source entered the community while infectious in NT^95^ | Community^97^ | Delta variant^97^ | No major restrictions^93,94^ | 3.222%^2,96^ |
| WA (Northern Suburbs) 27/06/2021 – 12/07/2021 (ongoing as at 30/06/2021) | 4 as at 30/06/2021^100,101^ | O | Epi link | 27/06/2021^99^ | 22/06/2021^99^ | Resident of Perth arrived back from NSW and tested positive on 27/06/2021^99^ | Source case travelled from NSW to WA, travelling around the community from 22/06/2021 - 24/06/2021^99^ | Community^99^ | Delta variant^99,101^ | Not reported | 1.471%^2^ |
| QLD  (NT Mine Workers Cluster) 27/06/2021 – 12/07/2021 (ongoing as at 30/06/2021) | 2 as at 30/06/2021 ^102^ | C | Epi link | 27/06/2021^103,105^ | 20/06/2021^104^ | Interstate FIFO miner who travelled from regional VIC to Brisbane where they stayed in a quarantine hotel and contracted virus on 18/06/2021, and then worked at the mine while infectious^102,107^ | Miner who flew into Brisbane travelled up to the Sunshine Coast tested positive after being informed on 26/06/2021 that co-worker at mine in NT had tested positive^104^ | Community^103^ | Delta variant^102^ | No major restrictions^106^ | 1.082%^2^ |
| QLD  (Prince Charles Hospital Cluster) 29/06/2021 – 13/07/2021 (ongoing as at 30/06/2021) | 2 as at 30/06/2021^110^ | O | Epi link | 29/06/2021^108^ | 19/06/2021^108^ | Not reported | Clerical assistant working immediately outside COVID-19 ward in the Prince Charles Hospital^108^ | Community^108^ | Delta variant^102,109^ | No major restrictions^106^ | 1.157%^2^ |

^*^When not reported publicly, ending dates of outbreaks/clusters are calculated as two weeks after the last case was reported

References

1) Government of South Australia. SA.GOV.AU:COVID-19. South Australia: Government of South Australia. 2020. [Available from: <https://www.covid-19.sa.gov.au/>.
2) covid19data.com.au. Infogram. 2021. Daily COVID-19 Tests Across Australia. [Available from: <https://infogram.com/1p7lyzng0l6j71fznrzp5jnqw6an669zy7y?live>.
3) Tomevska S. How did Adelaide's COVID-19 cluster begin and are medi-hotel procedures to blame?: ABC News; 17 November 2020. [Available from: <https://www.abc.net.au/news/2020-11-17/adelaide-coronavirus-cluster-how-it-began-explained/12891226>.
4) Gladstone N. In four days, 140,000 people across NSW have been tested for COVID-19: The Sydney Morning Herald; 23 Dec 2020. [Available from: <https://www.smh.com.au/national/nsw/in-four-days-140-000-people-across-nsw-have-been-tested-for-covid-19-20201223-p56pvk.html>.
5) Data.NSW. NSW Government. 2020. [Available from: <https://data.nsw.gov.au/data/dataset/nsw-covid-19-tests-by-location/resource/945c6204-272a-4cad-8e33-dde791f5059a>.
6) Hirst D. NSW Government to ease a raft of coronavirus restrictions in time for Christmas and New Year: ABC News; 25 Nov 2020. [Available from: https://www.abc.net.au/news/2020-11-25/nsw-government-to-ease-coronavirus-restrictions/12916818.
7) Cockburn P. New health rules apply to NSW residents in response to Northern Beaches COVID-19 cluster: ABC News; 19 Dec 2020. [Available from: <https://www.abc.net.au/news/2020-12-19/nsw-covid-rules-in-response-to-northern-beaches-cluster/12998408>.
8) NSW Government. Health. New South Wales. [Available from: <https://www.health.nsw.gov.au/> .
9) NSW Government. COVID-19 Weekly Surveillance in NSW: Summary for the week ending 2 January 2021. [www.health.nsw.gov.au/coronavirus. 7 Jan 2021](http://www.health.nsw.gov.au/coronavirus.%207%20Jan%202021). [Available from: https://www.health.nsw.gov.au/Infectious/covid-19/Documents/covid-19-surveillance-report-20210102.pdf.
10) NSW Government. COVID-19 (Coronavirus) Statistics. Health. 22 Dec 2020. [Available from: <https://www.health.nsw.gov.au/news/Pages/20201222_00.aspx>.
11) NSW Government. COVID-19 (Coronavirus) Statistics. Health. 30 Dec 2020. [Available from: <https://www.health.nsw.gov.au/news/Pages/20201230_00.aspx>.
12) Kidd J. Coronavirus-infected Sydney nurse linked to Avalon cluster not returning overseas travellers: ABC News; 24 Dec 2020. [Available from: <https://www.abc.net.au/news/2020-12-24/anglicare-nurse-in-avalon-cluster-not-connected-with-us-case/13012934> .
13) Cockburn P. What we know about the Croydon coronavirus cluster and what it means for Sydney: ABC News; 30 Dec 2020. [Available from: <https://www.abc.net.au/news/2020-12-30/what-we-know-about-croydon-covid-19cluster/13021302>.
14) Lathouris O. Four new local cases of coronavirus in NSW, people coming from Brisbane must self-isolate: 9News; 9 Jan 2021. [Available from: https://www.9news.com.au/national/coronavirus-nsw-health-update-11-new-cases-covid19-four-locally-acquired-latest-numbers/1b359362-0e2e-43a8-a5d3-fde4accc6801.
15) NSW Government. COVID-19 Restrictions for Greater Sydney. 20 Dec 2020. [Available from:<https://www.nsw.gov.au/media-releases/new-covid-19-restrictions-for-greater-sydney>.
16) Visontay E. Two Covid cases on NSW south coast linked to Melbourne cluster: The Guardian; 1 Jan 2021. [Available from: <https://www.theguardian.com/australia-news/2021/jan/01/two-covid-cases-on-nsw-south-coast-linked-to-melbourne-cluster>.
17) Cowie T, Sakkal P, Dow A. Two more COVID cases linked to Thai restaurant as testing wait times hit four hours: The Age; 1 Jan 2021. [Available from: <https://www.theage.com.au/national/victoria/covid-confirmed-in-regional-victoria-no-new-community-cases-20210101-p56r5e.html>.
18) Health and Human Services. Victoria State Government. 7 Jan 2021. Coronavirus update for Victoria – 7 January 2021. [Available from: <https://www.dhhs.vic.gov.au/coronavirus-update-victoria-7-january-2021>.
19) Woolley S. Genomic sequencing reveals new Sydney cluster is NOT linked to Northern Beaches: 7News; 3 Jan 2021. [Available from: <https://7news.com.au/lifestyle/health-wellbeing/genomic-sequencing-reveals-new-sydney-cluster-is-not-linked-to-northern-beaches-c-1882869>.
20) Lathouris O. Berala cluster: Sydney COVID outbreak linked to BWS where two infected staff worked throughout Christmas: 9News; 4 Jan 2021. [Available from: <https://www.9news.com.au/national/coronavirus-nsw-update-berala-bws-cluster-not-linked-to-avalon-sydney-northern-beaches-covid19-strain-genomic-testing-finds/26e4bfd0-df34-42d6-852c-5ac774bc8a11>.
21) NSW Government. COVID-19 (Coronavirus) Statistics. Health. 4 Jan 2021. [Available from: <https://www.health.nsw.gov.au/news/Pages/20210104_00.aspx>.
22) SBS News. Authorities on high alert as Melbourne mystery case attended MCG Boxing Day Test and Chadstone Shopping Centre: SBS News; 6 Jan 2021. [Available from: <https://www.sbs.com.au/news/authorities-on-high-alert-as-melbourne-mystery-case-attended-mcg-boxing-day-test-and-chadstone-shopping-centre>.
23) SBS News. Victoria's first mystery coronavirus case in two months linked to Sydney's Northern Beaches cluster: SBS News; 7 Jan 2021. [Available from: https://www.sbs.com.au/news/victoria-s-first-mystery-coronavirus-case-in-two-months-linked-to-sydney-s-northern-beaches-cluster.
24) Australian Bureau of Statistics. Brisbane and Perth have highest growth rates. ABS. 30 Mar 2021. [Available from: <https://www.abs.gov.au/media-centre/media-releases/brisbane-and-perth-have-highest-growth-rates>.
25) Stone L. How a casual cleaner’s unremarkable activities sparked a national health emergency: Brisbane Times; 13 Jan 2021. [Available from: <https://www.brisbanetimes.com.au/national/queensland/how-a-casual-cleaner-s-unremarkable-activities-sparked-a-national-health-emergency-20210111-p56t57.html>.
26) Queensland Health. Queensland borders unchanged. Queensland Government. 2 Jan 2021. [Available from: https://www.health.qld.gov.au/news-events/doh-media-releases/releases/queensland-borders-unchanged	.
27) Lynch L. Going viral: Timeline shows how COVID-19 infected Queensland: Brisbane Times; 27 Jan 2021. [Available from: <https://www.brisbanetimes.com.au/politics/queensland/going-viral-timeline-shows-how-covid-19-infected-queensland-20210112-p56tj7.html>
28) Premier and Minister for Trade. Queensland steps up border restrictions and re-emphasises safe behaviour. Queensland Government. 20 Dec 2020. [Available from: <https://statements.qld.gov.au/statements/91218>.
29) Queensland Health. Incident response set up following confirmation of Brisbane hotel cluster. Queensland Government. 13 Jan 2021. [Available from: <https://www.health.qld.gov.au/news-events/doh-media-releases/releases/incident-response-set-up-following-confirmation-of-brisbane-hotel-cluster>.
30) ABC News. Hotel quarantine worker tests positive for COVID-19 in Victoria, transmission between rooms being investigated: ABC News; 3 Feb 2021 [updated 4 Feb 2021. Available from: <https://www.abc.net.au/news/2021-02-03/victoria-investigating-possible-coronavirus-spread-in-quarantine/13117828>.
31) ABC News. Melbourne hotel quarantine worker who caught COVID is 'a model employee', Premier Daniel Andrews says: ABC News; 4 Feb 2021 [updated 5 Feb 2021. Available from: <https://www.abc.net.au/news/2021-02-04/vic-covid-case-at-hotel-quarantine-grand-hyatt-model-employee/13119826>.
32) Kinsella E. What we know about Victoria's new COVID-19 hotel quarantine cases: ABC News; 4 Feb 2021 [updated 4 Feb 2021. Available from: <https://www.abc.net.au/news/2021-02-04/how-covid-19-spread-in-two-victorian-quarantine-hotels/13118676>.
33) Wahlquist C. Australian Open hotel worker positive for Covid, with Victorian premier tightening restrictions: The Guardian; 4 Feb 2021. [Available from: <https://www.theguardian.com/australia-news/2021/feb/04/australian-open-hotel-quarantine-worker-positive-for-covid-with-victorian-premier-assuming-the-worst>.
34) Chapman A. Victorian hotel quarantine worker has UK strain of virus, CHO says: 7News; 5 Feb 2021 [updated 5 Feb 2021. Available from: <https://7news.com.au/lifestyle/health-wellbeing/victorian-hotel-quarantine-worker-has-uk-strain-of-virus-cho-says-c-2099036>.
35) ABC News. How the hotel quarantine outbreak that's landed Victorians in coronavirus lockdown spread: ABC News; 15 Feb 2021 [updated 15 Feb 2021. Available from: <https://www.abc.net.au/news/2021-02-15/how-did-victoria-outbreak-cases-escape-hotel-quarantine/13155644>.
36) Godde C. Vic Holiday Inn outbreak officially over: 7News; 14 Mar 2021. [Available from: <https://7news.com.au/lifestyle/health-wellbeing/vic-holiday-inn-outbreak-officially-over-c-2351570>.
37) Wahlquist C. 'Variant of concern': Victoria records five local Covid cases linked to hotel quarantine in past week: The Guardian; 9 Feb 2021. [Available from: <https://www.theguardian.com/australia-news/2021/feb/09/variant-of-concern-victoria-records-five-cases-of-coronavirus-in-hotel-quarantine-in-past-week>.
38) Michie F. Victoria is heading back into coronavirus lockdown tonight - These are the rules: ABC News; 12 Feb 2021 [updated 13 Feb 2021. Available from: <https://www.abc.net.au/news/2021-02-12/victoria-stage-4-lockdown-what-are-the-rules/13149252>.
39) Pearce L, Waters C. More exposure sites listed in Victoria as two new cases linked to Coburg venue: 9News; 14 Feb 2021. [Available from: <https://www.9news.com.au/national/coronavirus-victoria-new-exposure-sites-woolworths-aquatic-centre-swim-school-bakery-latest-updates/c36811e4-d846-497b-9f5f-d8ee036888b1>.
40) Ilanbey S. Quarantine worker’s false-negative test stalled discovery of Coburg exposure site: The Age; 14 Feb 2021. [Available from: <https://www.theage.com.au/national/victoria/quarantine-worker-s-false-negative-test-stalled-discovery-of-coburg-exposure-site-20210214-p572dj.html>.
41) Minister for Health and Ambulance Services. Renewed calls for mask wearing at airports. Queensland Government. 6 Mar 2021. [Available from: <https://statements.qld.gov.au/statements/91627>.
42) Robertson J. Inside Brisbane's COVID-19 crisis at the Princess Alexandra Hospital: ABC News; 3 Apr 2021 [updated 3 Apr 2021. Available from: <https://www.abc.net.au/news/2021-04-03/qld-covid-outbreak-princess-alexandra-hospital-lockdown-ppe/100045672>.
43) Vujkovic M. Eight new locally acquired COVID cases confirmed in Greater Brisbane linked to two clusters: ABC News; 30 Mar 2021 [updated 30 Mar 2021. Available from: https://www.abc.net.au/news/2021-03-30/queensland-coronavirus-greater-brisbane-lockdown-eight-cases/100035950 .
44) Gramenz E. Brisbane's COVID cluster grows as second nurse at PA hospital tests positive to coronavirus: ABC News; 31 Mar 2021 [updated 1 Apr 2021. Available from: <https://www.abc.net.au/news/2021-03-31/qld-covid-brisbane-lockdown-hospital-cluster/100034848> .
45) Queensland Health. Public Health Alert: PA Hospital. Queensland Government. 13 Mar 2021. [Available from: https://www.health.qld.gov.au/news-events/doh-media-releases/releases/public-health-alert-pa-hospital[.](https://www.health.qld.gov.au/news-events/doh-media-releases/releases/historic-case-linked-to-brisbane-covid-19-cluster)
46) Queensland Health. Historic case linked to Brisbane COVID-19 cluster. Queensland Government. 11 Mar 2021. [Available from: https://www.health.qld.gov.au/news-events/doh-media-releases/releases/historic-case-linked-to-brisbane-covid-19-cluster.
47) Motherwell S. How two clusters from one hospital triggered the Brisbane lockdown: ABC News; 30 Mar 2021 [updated 30 Mar 2021. Available from: <https://www.abc.net.au/news/2021-03-30/brisbane-lockdown-clusters-coronavirus-explained-pa-hospital/100037608>.
48) Perpitch N. Perth lockdown for three days will see Anzac Day services cancelled, residents urged to stay indoors: ABC News; 23 Apr 2021 [updated 23 Apr 2021. Available from: <https://www.abc.net.au/news/2021-04-23/perth-plunged-into-three-day-lockdown-after-hotel-covid/100091188>.
49) Department of the Premier and Cabinet. ANZAC Day 2021 dawn service. Government of Western Australia. 19 Apr 2021. [Available from: <https://www.wa.gov.au/government/announcements/anzac-day-2021-dawn-service>.
50) Department of Health. New community COVID-19 cases confirmed. Government of Western Australia. 1 May 2021. [Available from: <https://ww2.health.wa.gov.au/Media-releases/2021/New-community-COVID19-cases-confirmed>.
51) The Premier and Deputy Premier of Western Australia. Perth and Peel post-lockdown transition starts midnight tonight. Government of Western Australia. 26 Apr 2021. [Available from: <https://www.mediastatements.wa.gov.au/Pages/McGowan/2021/04/Perth-and-Peel-post-lockdown-transition-starts-midnight-tonight.aspx>.
52) De Poloni G. Perth hotel quarantine guard tests positive for COVID-19, along with two others: ABC News; 1 May 2021 [updated 2 May 2021. Available from: <https://www.abc.net.au/news/2021-05-01/perth-hotel-quarantine-guard-tests-positive-for-covid-19/100109788>.
53) Fiore B. Coronavirus: WA records no new cases of COVID-19 as restrictions ease across Perth and Peel regions: PerthNow; 8 May 2021. [Available from: <https://www.perthnow.com.au/news/coronavirus/coronavirus-wa-records-no-new-cases-of-covid-19-as-restrictions-ease-across-perth-and-peel-regions-ng-b881867151z>.
54) NSW Government. COVID-19 (Coronavirus) Statistics. Health. 17 May 2021. [Available from: <https://www.health.nsw.gov.au/news/Pages/20210517_00.aspx>.
55) NSW Government. Public health alert - COVID-19 case. Health. 5 May 2021. [Available from: <https://www.health.nsw.gov.au/news/Pages/20210505_01.aspx>.
56) Woolley S. NSW Health reveals likely source of local COVID-19 infections as new restrictions are rolled out: 7News; 6 May 2021 [updated 6 May 2021. Available from: <https://7news.com.au/lifestyle/health-wellbeing/nsw-health-reveals-likely-source-of-local-covid-19-infections-as-new-restrictions-are-rolled-out-c-2771446>.
57) NSW Government. COVID-19 (Coronavirus) Statistics. Health. 3 May 2021. [Available from: <https://www.health.nsw.gov.au/news/Pages/20210503_00.aspx>.
58) Wahlquist C. Genomic sequencing under way after four people in Melbourne test positive: The Guardian; 25 May 2021. [Available from: <https://www.theguardian.com/australia-news/2021/may/24/two-people-in-melbournes-northern-suburbs-likely-test-positive-to-covid>.
59) Davey M. Victorian authorities blame new Covid case on South Australian hotel quarantine: The Guardian; 11 May 2021. [Available from: <https://www.theguardian.com/australia-news/2021/may/11/victorian-man-tests-positive-to-covid-after-completing-hotel-quarantine-in-south-australia>.
60) Health and Human Services. Victoria State Government. 9 June 2021. Coronavirus update for Victoria - 9 June 2021. [Available from: <https://www.dhhs.vic.gov.au/coronavirus-update-victoria-9-june-2021>.
61) Boseley M. Melbourne aged care resident and nurse test positive for Covid as Victoria records four new cases: The Guardian; 6 June 2021. [Available from: <https://www.theguardian.com/australia-news/2021/jun/06/melbourne-aged-care-resident-and-nurse-test-positive-for-covid-as-victorias-records-four-new-cases>.
62) Cunningham M, Preiss B, Cowie T. What we know so far about Melbourne’s new coronavirus cases: The Sydney Morning Herald; 25 May 2021 [updated 26 May 2021. Available from: <https://www.smh.com.au/national/what-we-know-so-far-about-melbourne-s-new-coronavirus-cases-20210525-p57uw8.html>
63) Health and Human Services. Media Hub—coronavirus (COVID-19): Victoria State Government [updated 23 Nov 2021. Available from: <https://www.dhhs.vic.gov.au/media-hub-coronavirus-disease-covid-19>.
64) 7News. How Melbourne's virus outbreak has spread: 7News; 26 May 2021 [updated 26 May 2021. Available from: <https://7news.com.au/lifestyle/health-wellbeing/how-melbournes-virus-outbreak-has-spread-c-2936054>
65) Health and Human Services. Victoria State Government. 26 May 2021. Coronavirus update for Victoria – 26 May 2021. [Available from: <https://www.dhhs.vic.gov.au/coronavirus-update-victoria-26-may-2021>.
66) Health and Human Services. Victoria State Government. 19 June 2021. Coronavirus update for Victoria – 19 June 2021. [Available from: <https://www.dhhs.vic.gov.au/coronavirus-update-victoria-19-june-2021>.
67) Risso A, Godde C. Worker case locks down Vic aged care home: 7News; 30 May 2021 [updated 30 May 2021. Available from: <https://7news.com.au/lifestyle/health-wellbeing/five-new-local-vic-cases-amid-lockdown-c-2968310>.
68) Willingham R. Victoria enters COVID-19 lockdown as cases from Melbourne outbreak grow: ABC News; 27 May 2021 [updated 28 May 2021. Available from: <https://www.abc.net.au/news/2021-05-27/victoria-covid-cases-melbourne-outbreak-lockdown-restrictions/100169172>.
69) ABC News. Victoria's nine new COVID cases all linked to existing outbreaks, include three children: ABC News; 7 June 2021 [updated 7 June 2021. Available from: <https://www.abc.net.au/news/2021-06-07/victoria-records-new-covid-cases-as-lockdown-deadline-looms/100194240>.
70) ABC News. Delta COVID-19 strain that devastated India detected in Victorian outbreak: ABC News; 4 June 2021 [updated 4 June 2021. Available from: <https://www.abc.net.au/news/2021-06-04/vic-delta-variant-of-coronavirus-detected-in-australia/100190460>.
71) Dunstan J. How the Delta COVID variant likely jumped Victorian hotel quarantine and started a Melbourne outbreak: ABC News; 8 June 2021 [updated 9 June 2021. Available from: <https://www.abc.net.au/news/2021-06-08/melbourne-covid-outbreak-delta-strain-link-hotel-quarantine/100183468>.
72) ABC News. Victoria records no new local COVID cases as Melbourne emerges from two-week lockdown: ABC News; 11 June 2021 [updated 11 June 2021. Available from: <https://www.abc.net.au/news/2021-06-11/victoria-new-covid-cases-melbourne-lockdown-lifts/100207318>.
73) Health and Human Services. Victoria State Government. 10 June 2021. Coronavirus update for Victoria – 10 June 2021. [Available from: <https://www.dhhs.vic.gov.au/coronavirus-update-victoria-10-june-2021>
74) ABC News. Here are the new COVID restrictions for regional Victoria and Melbourne: ABC News; 3 June 2021 [updated 16 Sept 2021. Available from: <https://www.abc.net.au/news/2021-06-03/victoria-lockdown-restrictions-regional-and-greater-melbourne/100186702>.
75) ABC News. List of COVID-19 exposure sites for Queensland includes Caloundra cafe and Bunnings: ABC News; 9 June 2021 [updated 12 June 2021. Available from: <https://www.abc.net.au/news/2021-06-09/queensland-covid-exposure-sites-list-announced-positive-case/100202484>.
76) Riga R. Victorian couple who tested positive to COVID fined $4k each over alleged border breaches: ABC News; 18 June 2021 [updated 19 June 2021. Available from: <https://www.abc.net.au/news/2021-06-18/queensland-covid-couple-fined-border-breaches-hotspot/100223284>
77) ABC News. Victoria moves to prevent COVID-19 outbreak at Melbourne Southbank townhouse complex: ABC News; 14 June 2021 [updated 14 June 2021. Available from: <https://www.abc.net.au/news/2021-06-14/victoria-new-covid-cases-melbourne-restrictions-on-track-to-ease/100212806>.
78) Health and Human Services. Victoria State Government. 26 June 2021. Coronavirus update for Victoria – 26 June 2021. [Available from: https://www.dhhs.vic.gov.au/coronavirus-update-victoria-26-june-2021.
79) Health and Human Services. Victoria State Government. 12 June 2021. Coronavirus update for Victoria – 12 June 2021. [Available from: <https://www.dhhs.vic.gov.au/coronavirus-update-victoria-12-june-2021>.
80) ABC News. Victoria records two new local COVID-19 cases as authorities fight Southbank apartment complex outbreak: ABC News; 15 June 2021 [updated 16 June 2021. Available from: <https://www.abc.net.au/news/2021-06-15/victoria-records-two-new-cases-of-covid-19-southbank-outbreak/100215184>
81) NSW Government. COVID-19 in NSW - up to 8pm 29 November 2021 [Available from: https://www.health.nsw.gov.au/Infectious/covid-19/Pages/stats-nsw.aspx#weekly.
82) Ursula M. How the potentially 'inexcusable' actions of a limo driver put Sydney on COVID-19 alert: ABC News; 2021 [updated 18 Jun 2021. Available from: https://www.abc.net.au/news/2021-06-17/nsw-quarantine-worker-may-have-breached-health-order/100223120.
83) McGowan M. Police probe into Sydney limousine driver expanded as health minister seeks tougher mask rules: The Guardian; 2021 [updated 24 Jun 2021. Available from: https://www.theguardian.com/australia-news/2021/jun/24/police-probe-into-sydney-limousine-driver-expanded-as-health-minister-seeks-tougher-mask-rules.
84) Australian Bureau of Statistics. 2016 Census QuickStats, Zetland [updated 26 October 2021. Available from: https://quickstats.censusdata.abs.gov.au/census_services/getproduct/census/2016/quickstat/SSC14524?opendocument.
85) Daoud E. Sydney’s COVID cluster grows as new cases confirmed: 7 News; 2021 [updated 17 Jun 2021. Available from: https://7news.com.au/travel/coronavirus/sydneys-covid-cluster-grows-as-new-cases-confirmed--c-3136422.
86) NSW Government. COVID-19 rules [updated 24 November 2021. Available from: https://www.nsw.gov.au/covid-19/stay-safe/rules.
87) Taouk M. From Bondi to Wollongong: Tracking Sydney's latest COVID-19 outbreak: ABC News; 23 Jun 2021 [updated 20 Aug 2021. Available from: https://www.abc.net.au/news/2021-06-23/nsw-covid-bondi-outbreak-timeline/100237834.
88) King J. Queensland authorities trace source of Emirates passenger's COVID-19 infection after ruling out Delta variant ABC News; 21 Jun 2021 [updated 21 Jun 2021. Available from: https://www.abc.net.au/news/2021-06-21/traveller-covid-19-ruling-out-delta-variant/100226752.
89) Nothling L. Queensland records three locally acquired COVID-19 cases linked to flight attendant: ABC News; 24 Jun 2021 [updated 24 Jun 2021. Available from: https://www.abc.net.au/news/2021-06-24/covid-qld-case-portuguese-family-centre-tests-positive/100239866.
90) Edwards N, Masters R. Queensland introduces new border pass system: 9 News; 17 Jun 2021 [Available from: https://www.9news.com.au/national/queensland-announced-new-border-pass-along-with-sydney-melbourne-restrictions/ffac916f-0492-4d32-8cd2-1b2f177ec50d.
91) Queensland Government. Brisbane’s COVID-19 community case update – variant confirmed 21 Jun 2021 [Available from: https://www.health.qld.gov.au/news-events/doh-media-releases/releases/brisbanes-covid-19-community-case-update-variant-confirmed.
92) Purtell M. Queensland records two new locally acquired COVID-19 cases linked to Portuguese restaurant: ABC News; 25 Jun 2021 [Available from: https://www.abc.net.au/news/2021-06-25/queensland-records-two-new-locally-acquired-covid-19-cases/100226750.
93) Thompson J. Darwin region sent into snap lockdown with five new Granites mine COVID-19 cases, fears more to come 27 Jun 2021 [Available from: https://www.abc.net.au/news/2021-06-27/darwin-into-snap-lockdown-nt-four-coronavirus-cases/100247354.
94) Heaney C. Northern Territory declares Greater Sydney, Blue Mountains and Wollongong COVID-19 hotspots: ABC News; 23 Jun 2021 [Available from: https://www.abc.net.au/news/2021-06-23/nt-hotspot-declaration-nsw-quarantine-sydney/100237390.
95) Northern Territory Government. Positive COVID-19 Case Update coronavirus.nt.gov.au 26 June 2021 [Available from: https://coronavirus.nt.gov.au/updates/items/2021-06-26-positive-covid-19-case-update.
96) Northern Territory Government. Population: Northern Territory Government Department of Industry, Tourism and Trade,; 2021 [Available from: https://industry.nt.gov.au/economic-data-and-statistics/business/business-statistics/nt-key-business-statistics/population.
97) Saunokonoko M. NT gold mine worker tests COVID-19 positive, putting 1600 others at risk: 9 News; 26 Jun 2021 [Available from: https://www.9news.com.au/national/nt-granites-gold-mine-worker-in-tanami-desert-tests-positive-covid-19/2f101812-117f-44a6-ad5c-2b6a1c76dc72.
98) Northern Territory Government. Northern Territory Media Release 28 June 2021 [Available from: https://mediareleases.nt.gov.au/article?id=34624.
99) ABC News. Perth and Peel are in lockdown. Here's what you need to know about the new rules 29 Jun 2021 [updated 1 Jul 2021. Available from: https://www.abc.net.au/news/2021-06-29/new-rules-for-perth-peel-lockdown-masks-schools-gyms/100250936.
100) Shine R. One new WA COVID case on second day of Perth lockdown sparked by Delta coronavirus variant: ABC News; 30 Jun 2021 [Available from: https://www.abc.net.au/news/2021-06-30/one-new-wa-covid-case-on-day-two-of-perth-coronavirus-lockdown/100254474.
101) de Kruijff P. WA records no new COVID-19 cases, buoying hopes lockdown will lift: WAtoday; July 1, 2021 [Available from: https://www.watoday.com.au/national/western-australia/wa-records-no-new-covid-19-cases-buoying-hopes-lockdown-will-lift-20210701-p585y7.html.
102) Purtell M, McKenna K. Queensland has four COVID clusters — three of them Delta variant. This is what we know so far: ABC News; 1 Jul 2021 [Available from: https://www.abc.net.au/news/2021-07-01/queensland-coronavirus-clusters-three-delta-explainer/100255826.
103) Cramsie E. Three new COVID cases recorded in Queensland, including two detected in the community: ABC News; 28 Jun 2021 [Available from: https://www.abc.net.au/news/2021-06-28/queensland-coronavirus-three-new-cases-two-in-community/100248558.
104) Saunokonoko M. More restrictions, mask mandate announced for Queensland as female miner confirmed to have Delta strain: 9News; 28 June 2021 [Available from: https://www.9news.com.au/national/queensland-announces-new-covid-restrictions-as-female-miner-confirmed-to-have-delta-

strain/83c408a3-7b8e-4ec0-b83f-e4b5d715b487.

105) Queensland Government. Further COVID-19 case identified in Queensland 27 Jun 2021 [Available from: https://www.health.qld.gov.au/news-events/doh-media-releases/releases/further-covid-19-case-identified-in-queensland.
106) Queensland Government. QLD responds swiftly to NSW’s lockdown 25 Jun 2021 [Available from: https://www.health.qld.gov.au/news-events/doh-media-releases/releases/qld-responds-swiftly-to-nsws-lockdown2.
107) Smee B. NT miner may have caught Covid from air conditioning at Brisbane quarantine hotel: The Guardian; 7 Jul 2021 [Available from: https://www.theguardian.com/australia-news/2021/jul/07/nt-miner-may-have-caught-covid-from-air-conditioning-at-brisbane-quarantine-hotel.
108) Ruddick B. Queenslanders heading for three-day lockdown after unvaccinated hospital worker tests positive to COVID-19: ABC News; 29 Jun 2021 [Available from: https://www.abc.net.au/news/2021-06-29/queensland-coronavirus-delta-palaszczuk-lockdown/100249626.
109) Queensland Government. Queensland COVID-19 update 29 June 2021 [Available from: https://www.health.qld.gov.au/news-events/doh-media-releases/releases/queensland-covid-19-update9.
110) Read C, Dennien M. Infected Brisbane hospital receptionist did not ‘take option up’ to get vaccinated: Brisbane Times; 30 Jun 2021 [Available from: <https://www.brisbanetimes.com.au/national/queensland/infected-brisbane-hospital-receptionist-did-not-take-option-up-to-get-vaccinated-20210630-p585g8.html>.
